# Modeling infections and deaths averted due to COVID-19 vaccination strategies in Ghana

**DOI:** 10.1101/2022.07.09.22277458

**Authors:** Sylvia K. Ofori, Jessica S. Schwind, Kelly L. Sullivan, Gerardo Chowell, Benjamin J. Cowling, Isaac Chun-Hai Fung

## Abstract

This study assessed the impact of various COVID-19 vaccination strategies on health outcomes in Ghana using an age-stratified compartmental model. The population was stratified into three age groups: <25 years, 25-64 years, and 65+ years. Five vaccination optimization scenarios were explored, assuming that one million persons could be vaccinated in three versus six months. We also performed uncertainty analysis by assuming that the available doses were halved and doubled. The vaccine optimization strategies were assessed for the initial strain, followed by a sensitivity analysis for the delta variant by varying the reproduction number and vaccine efficacy. The results showed that vaccinating individuals <65 years was associated with the lowest cumulative infections when one million persons were vaccinated over three months for both the initial strain and the delta variant. On the contrary, prioritizing the elderly (65+) was associated with the lowest cumulative deaths for both strains.

**One-sentence summary:** An age-stratified model of COVID-19 vaccination in Ghana found vaccinating individuals <65 years was associated with the lowest cumulative infections when one million persons were vaccinated over three months while prioritizing the elderly (65+) was associated with the lowest cumulative deaths.

## Introduction

Since the first case of coronavirus disease 2019 (COVID-19), caused by severe acute respiratory syndrome coronavirus 2 (SARS-CoV-2), was reported in China (*1*), the pandemic has spread to Africa (*2*). Ghana reported its first case on March 12, 2020, with 161,157 cases and 1,445 deaths recorded as of April 21, 2022,(*3, 4*). The country introduced various public health measures when the pandemic first hit the country, including school closures, travel bans, mask mandates, and vaccination, which were associated with a decline in transmission (*5*). Ghana was the first country to receive 600,000 doses of the Oxford-AstraZeneca COVID-19 vaccine AZD1222 (brand name: Vaxzevria) vaccine on February 24, 2021, through the COVAX program (*6*). The vaccination program was deployed in March 2021, with politicians and civil society leaders publicly taking shots to boost the trust in the vaccine program (*7*). The first batch of the vaccines was targeted at the regions with the highest burden of COVID-19, namely, the Greater Accra and Ashanti regions, frontline healthcare, the elderly, and persons with comorbidities. In addition to the initial doses, the ministry of health received an additional supply of the AZD1222 vaccine and the Pfizer-BioNTech BNT162b2 (brand name: Tozinameran) vaccine from several high-income countries, including Germany, Denmark, and Norway (*8*).

Due to the limited availability of doses and vaccine hesitancy, only 9.2% of the Ghanaian population of 30,800,000 was fully vaccinated as of December 2021 (*9*). Hence, the government’s goal to reach widespread vaccine coverage by October 2021 was not met (*6*). Studies on COVID- 19 vaccine hesitancy among Ghanaians reported that more than 35% of participants said they would not receive the vaccine because of concerns about vaccine efficacy and conspiracy theories (*7, 10*). Moreover, a seroprevalence study found that 19% of Ghanaians tested positive to anti-SARS-CoV-2 IgM or IgG or both in August 2020 (*11*). Thus, a majority of Ghanaians may be susceptible, and insight to effectively prioritize dispensation of limited vaccines would be the best approach to mitigating the pandemic. To optimize Ghana’s vaccination strategy and provide evidence of the benefits of vaccination and increase uptake in the population, we need to quantify the vaccine’s impact on the magnitude of the epidemic peak, cumulative infections, and deaths due to limited vaccine supplies and logistical reasons.

Several mathematical modeling studies on COVID-19 vaccination strategies in other jurisdictions were published in 2020-21 (*12-18*). Alagoz and colleagues used an agent-based model to simulate the transmission dynamics of COVID-19, accounting for the proportion of the population vaccinated, vaccine capacity, and adherence to nonpharmaceutical interventions (*12*). Moghadas et al. used an agent-based model to evaluate the impact of vaccination campaigns on reducing the incidence, hospitalizations, and death (*15*). Aside from agent-based models, homogenous-mixing and age-stratified compartmental models have also been used. Matrajt et al. used an age-stratified deterministic model paired with optimization algorithms for sixteen age groups by varying vaccination efficacy and coverages in the population (*16*). Mumtaz and colleagues used an age-stratified model to assess the vaccination rollout under different vaccination coverages accounting for the decline in transmission and age-mixing matrix (*17*). Bubar et al. expanded their work further to account for contact structure, seroprevalence, and age-specific vaccine efficacy (*18*). The outcomes explored in these studies include symptomatic infections, cumulative infections and deaths, and hospitalizations, focusing mainly on high-income countries outside Africa. Thus, the question of who to vaccinate first when vaccines are available, and the sensitivity of modeling outputs to the choice of contact matrices are under-explored in the African context and Ghana specifically.

The purpose of this study is to use an age-stratified model to assess the impact of vaccinating one million persons in three versus six months using two African contact matrices. We retrospectively assessed the counterfactual impact of various age-targeted vaccine optimization strategies against the initial and delta strains of SARS-CoV-2 when vaccines first became available. The study’s findings would facilitate future vaccination programs by identifying relevant factors to be considered to achieve the best outcomes.

## Methods

### Model formulation

We proposed an age-stratified Susceptible-Exposed-Presymptomatic-Symptomatic-Asymptomatic-Recovered-Dead-Vaccinated (SEPIARD-V) model to simulate SARS-CoV-2 transmission dynamics and the impact of various vaccination scenarios (Figure S1 in Technical Appendix A) (*19*). The SEPIARD-V model acknowledges that individuals who are initially asymptomatic and later develop symptoms transmit the virus while in the presymptomatic phase. Individual studies included in a systematic review on the transmission dynamics provided evidence of the presymptomatic transmission of COVID-19 1-3 d before symptom onset (*20*). The model was suitable for studying the transmission dynamics of COVID-19 in Ghana due to the growing evidence that both symptomatic and asymptomatic patients transmit the infection regardless of their symptomatic status (*21-23*). Our model, therefore, assumed that presymptomatic, asymptomatic, and symptomatic individuals contributed to transmission. The model also assumed that immunity from both natural infection and vaccination waned over time, making reinfection possible (see Table 1 and Technical Appendix A for model parameters). A basic reproduction number (R0) of 3.13 was assumed for the virus in the main analysis to represent the virus strain that first hit Ghana in spring of 2020, referred to as the “initial strain” in this study (*24*). A higher R_0_ at 5.35 was assumed for the delta variant in the scenario analyses with a reduced vaccine efficacy of 67% for the AZD1222 vaccine (*25, 26*).

**Table 1:** Parameter values for the age-stratified SEPIARD-V COVID-19 model

| Parameter | Symbol | Value | Reference |
| --- | --- | --- | --- |
| Mean latency period which is the period from exposure to infectiousness | $1/k$ | 1.85 d | Abbasi et al., (43), Liu et al. (44) |
| Mean duration of being infectious and symptomatic | $1/f$ | 15.7 d | Cai et al., (45) Xing et al., (46) |
| Mean duration of being infectious and asymptomatic | $1/q$ | 7.25 d | Ma et al., (47), Byrne et al. (48) |
| Mean duration of being infectious and presymptomatic | $1/c$ | 2.9 d | Tindale et al.,(49), Byrne et al.,(48) |
| Reproduction number for the initial strain | $R_0$ | 3.13 | Armachie et al., (24) |
| Reproduction number for the delta strain | $R_0$ | 5.35 | Pearson et al., (25) |
| Probability of exposed person becoming pre-symptomatically infected | $\delta$ | 0.30 | Chen et al., (50) Buitrago-Garcia et al., (51) |
| Vaccine efficacy against infection | $\sigma$ | 0.745 | Knoll et al., (52) |
| Relative transmissibility of asymptomatic individuals | $u$ | 0.75 | CDC (53) |
| Relative transmissibility of presymptomatic individuals | $r$ | 0.75 | CDC (53) |
| Mean duration of immunity following vaccination | $\chi$ | 182 d | Iacobucci (54) |
| Mean duration of immunity following natural infection | $w$ | 365 d | Good & Hawke (55) |
| Age-specific case fatality ratio | $z$ | 0.002 for <25 y, 0.005 for 25-64 y, 0.048P for 65+ | Our world in Data (Range from January 26, 2021, and November 12, 2021), Yakubu(56) |
| Daily vaccination rate | $v$ | Varied between 0.00009 - 0.0163977 d<sup>-1</sup> per person | Estimated |

The model was run for 500 d to allow enough time for the first wave of the epidemic to subside and observe when the second wave began to emerge.

### Age groups and contact matrices

Due to the strong evidence of assortative mixing between age groups in Sub-Saharan Africa (*27, 28*), a contact matrix between age groups was incorporated into the model. The population was stratified into three groups: <25 y, 25-64 y, and 65+ y, and two contact matrices were adapted from studies in Uganda (“main matrix”) and Ethiopia (“second matrix”) (*27, 29*). Briefly, the “main matrix” suggested that, on average, the within-group contact rate among individuals below 25 y was 23.58 per day; for those between 25-64 y, it was 15.05 per day; and it was 0.54 per day for those above 64 y (*27*). For the second matrix, on average, the within-group daily contact rate among individuals below 25 y was 8.2; for those between 25-64 y, it was 7.8; and it was 1.6 for those above 64 y (*29*). The model was parameterized to the Ghanaian population, with 56.08% <25 y (n=17,248,000), and 4.44% 65+ y (n=1,367,520) (*30*). See Technical Appendix A for details.

### Scenario analyses

This analysis aimed to determine which age group should be prioritized in the case of limited vaccine supply under different rollout speeds. We analyzed the following scenarios of the percentage of each sub-population vaccinated when prioritizing different age groups, with coverage calculated for one million people using the 2021 population (*31*): (i) 73.8% of the 65+ y, (ii) 8.2% of those between 25-64 y, (iii) 5.8% of persons below 25 y, and (iv) 3.4% of persons below 65 y would be vaccinated in each sub group; (v) we also assessed the impact of vaccinating each age group at the same rate without prioritization. Two rollout speeds (daily vaccination rates) were used in each scenario, assuming two million doses can be exhausted in three months and six months. Details of the daily vaccination rates (*v*) for each scenario can be found in Tables S1-S3 in Technical Appendix A.

Finally, we performed analysis for two additional scenarios by changing the assumptions on vaccine supplies. First, the number of people to be vaccinated was either halved or doubled. Hence, we assumed enough vaccines were available for 500,000 (one million doses) and 2,000,000 (four million doses) people. The scenario analysis described above was repeated using the second contact matrix adapted from Trentini et al. (*29*).

### Analysis

The model’s system of ordinary differential equations was solved following the Runge-Kutta 4 method in the deSolve package in R version 4.1.1 (R Core Team, R Foundation for Statistical Computing, Vienna, Austria) (*32*). The number of infections and deaths averted in the general population was estimated and compared across the aforementioned scenarios. Furthermore, the percent of the population who were symptomatic at the peak, ever infected (cumulative infections), and cumulative deaths were assessed. See Technical Appendix A for methodological details and Technical Appendix B for the R code.

### Ethics

The Georgia Southern University Institutional Review Board determined that this project (H20364) was exempt from full review under the non-human subjects determination (G8) according to the Code of Federal Regulations Title 45 Part 46.

## Results

### Symptomatic infections at the peak under the main scenario of vaccinating one million persons

The following results of our main analysis assumed an R_0_ of 3.13 for the initial strain. The results showed that vaccinating one million individuals <65 years in 3 months were associated with the lowest percentage (6.78%) of symptomatic individuals in the population at the peak. However, prioritizing the elderly resulted in the highest percentage of symptomatic individuals (7.19%) at the peak, given a three-month rollout using the main matrix. Similar results were obtained with the second matrix. If the rollout period was increased to six months, prioritizing persons <25 years had the lowest symptomatic percentages (7.01%) using the main matrix, while the second matrix suggested focusing on 25-64 years was associated with the lowest symptomatic infections (6.96%) (Figure 1; Tables 2 and 3).

**Figure 1:**
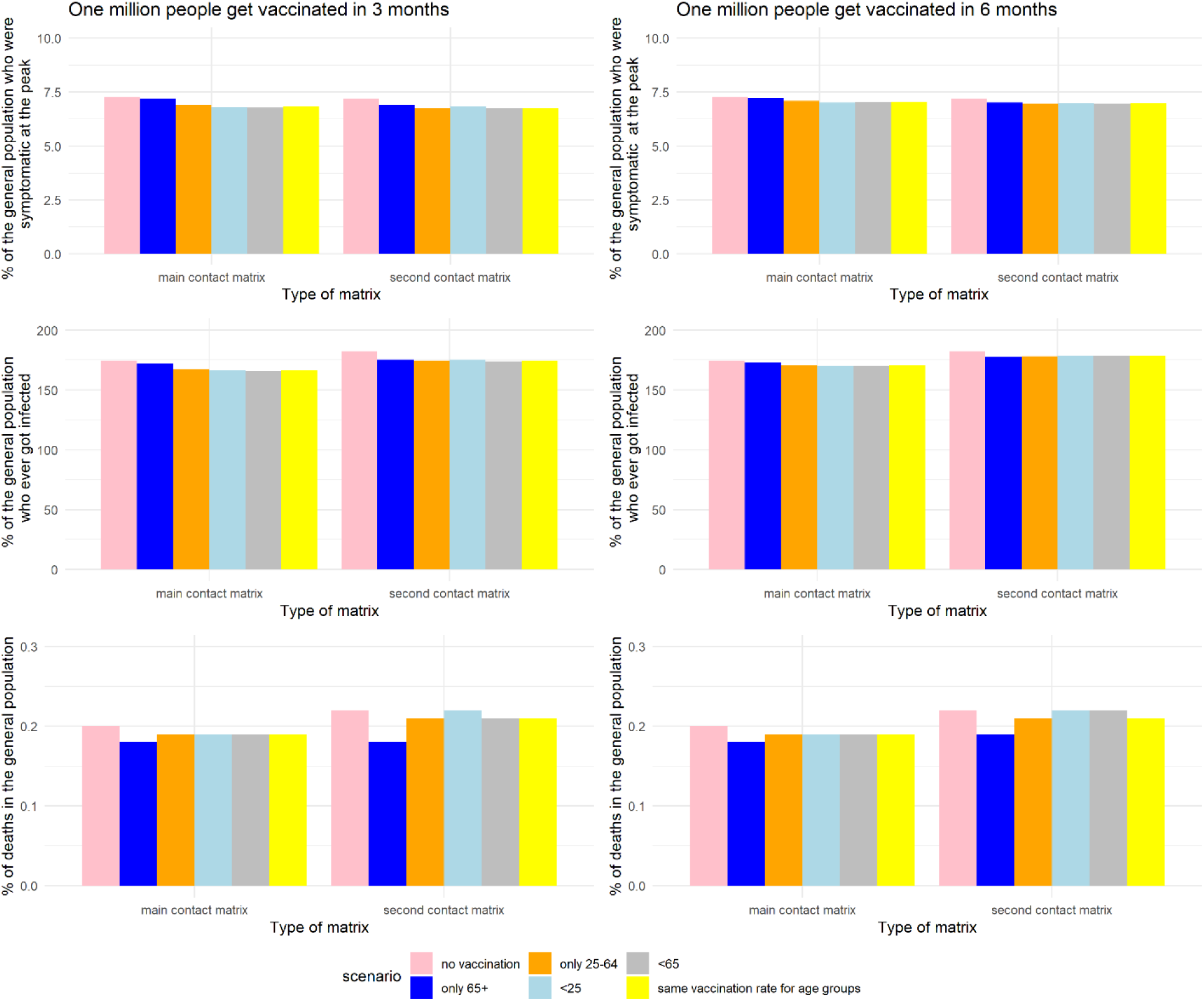
The impact of various vaccination scenarios on symptomatic infections at peak (upper panel), cumulative infections (middle panel), and deaths (lower panel) as a percentage of the general population with two different contact matrices, in the main analysis with R0=3.13 (initial strain). *% of cumulative infections is greater than 100% because of waning immunity from natural infection and vaccination

**Table 2:** Scenario analysis of outcomes in the total population under various vaccination scenarios using the main matrix for the initial strain

| **Vaccine prioritization** | **500,000 people vaccinated in 3 months (%)** | **500,000 people vaccinated in 6 months (%)** | **One million people vaccinated in 3 months (%)** | **One million people vaccinated in 6 months (%)** | **Two million people were vaccinated in 3 months (%)** | **Two million people were vaccinated in 6 months (%)** |
| --- | --- | --- | --- | --- | --- | --- |
| **Symptomatic infections at peak** |  |  |  |  |  |  |
| Only 65+ | 7.22 | 7.24 | 7.19 | 7.22 | 7.16 | 7.19 |
| 25-64 | 7.09 | 7.17 | 6.92 | 7.09 | 6.61 | 6.92 |
| <25 | 7.01 | 7.13 | 6.79 | 7.01 | 6.25 | 6.75 |
| < 65 | 7.03 | 7.15 | 6.78 | 7.03 | 6.37 | 6.81 |
| Same vaccination rate across age groups | 7.04 | 7.15 | 6.83 | 7.04 | 6.40 | 6.83 |
| **Cumulative infections** |  |  |  |  |  |  |
| Only 65+ | 172.87 | 173.47 | 172.08 | 172.87 | 171.35 | 172.08 |
| 25-64 | 170.80 | 172.57 | 167.51 | 170.89 | 161.20 | 167.44 |
| <25 | 170.04 | 172.20 | 166.29 | 170.04 | 157.04 | 165.76 |
| < 65 | 170.28 | 172.43 | 165.76 | 170.28 | 158.20 | 166.19 |
| Same vaccination rate across age groups | 170.41 | 172.39 | 166.42 | 170.41 | 158.47 | 166.42 |
| **Deaths** |  |  |  |  |  |  |
| Only 65+ | 0.18 | 0.19 | 0.18 | 0.18 | 0.17 | 0.17 |
| 25-64 | 0.19 | 0.19 | 0.19 | 0.19 | 0.18 | 0.19 |
| <25 | 0.19 | 0.19 | 0.19 | 0.19 | 0.18 | 0.19 |
| < 65 | 0.19 | 0.19 | 0.19 | 0.19 | 0.18 | 0.19 |
| Same vaccination rate across age groups | 0.19 | 0.19 | 0.19 | 0.19 | 0.18 | 0.19 |

**Table 3:** Scenario analysis of outcomes in the total population under various vaccination scenarios using the second matrix for the initial strain

| **Vaccine prioritization** | **500,000 people vaccinated in 3 months (%)** | **500,000 people vaccinated in 6 months (%)** | **One million people vaccinated in 3 months (%)** | **One million people vaccinated in 6 months (%)** | **Two million people were vaccinated in 3 months (%)** | **Two million people were vaccinated in 6 months (%)** |
| --- | --- | --- | --- | --- | --- | --- |
| **Symptomatic infections at peak** |  |  |  |  |  |  |
| Only 65+ | 7.02 | 7.09 | 6.92 | 7.02 | 6.82 | 6.92 |
| 25-64 | 6.96 | 7.07 | 6.76 | 6.96 | 6.36 | 6.75 |
| <25 | 6.99 | 7.09 | 6.82 | 6.98 | 6.41 | 6.79 |
| < 65 | 6.97 | 7.08 | 6.74 | 6.97 | 6.35 | 6.76 |
| Same vaccination rate across age groups | 6.98 | 7.08 | 6.76 | 6.98 | 6.35 | 6.76 |
| **Cumulative infections** |  |  |  |  |  |  |
| Only 65+ | 177.68 | 179.54 | 175.32 | 177.68 | 173.23 | 175.33 |
| 25-64 | 178.22 | 180.28 | 174.34 | 178.22 | 166.67 | 174.26 |
| <25 | 178.62 | 180.48 | 175.41 | 178.62 | 167.63 | 174.95 |
| < 65 | 178.39 | 180.48 | 174.00 | 178.39 | 166.69 | 174.42 |
| Same vaccination rate across age groups | 178.30 | 180.33 | 174.23 | 178.30 | 166.10 | 174.23 |
| **Deaths** |  |  |  |  |  |  |
| Only 65+ | 0.19 | 0.20 | 0.18 | 0.19 | 0.17 | 0.18 |
| 25-64 | 0.21 | 0.22 | 0.21 | 0.21 | 0.19 | 0.21 |
| <25 | 0.22 | 0.22 | 0.22 | 0.22 | 0.21 | 0.21 |
| < 65 | 0.22 | 0.22 | 0.21 | 0.22 | 0.20 | 0.21 |
| Same vaccination rate across age groups | 0.21 | 0.22 | 0.21 | 0.21 | 0.20 | 0.21 |

### Cumulative infections under the main scenario of vaccinating one million persons

Our results suggest that vaccinating persons <65 years was associated with the largest number of cumulative infections averted in Ghana under the assumption of vaccinating one million people in three months (2,652,484 cases), whereas vaccinating persons 65+ years was associated with the smallest number averted (705,981 cases) (Figure 2). We also found that vaccinating persons <25 years should be prioritized when the population was vaccinated at a slower rate (over six months) or when the vaccine supply doubled or halved (Table 2). While the results for the period of three months are robust to a change in the contact matrix, a more extended period (six months) seemed to be sensitive (Table 3).

**Figure 2:**
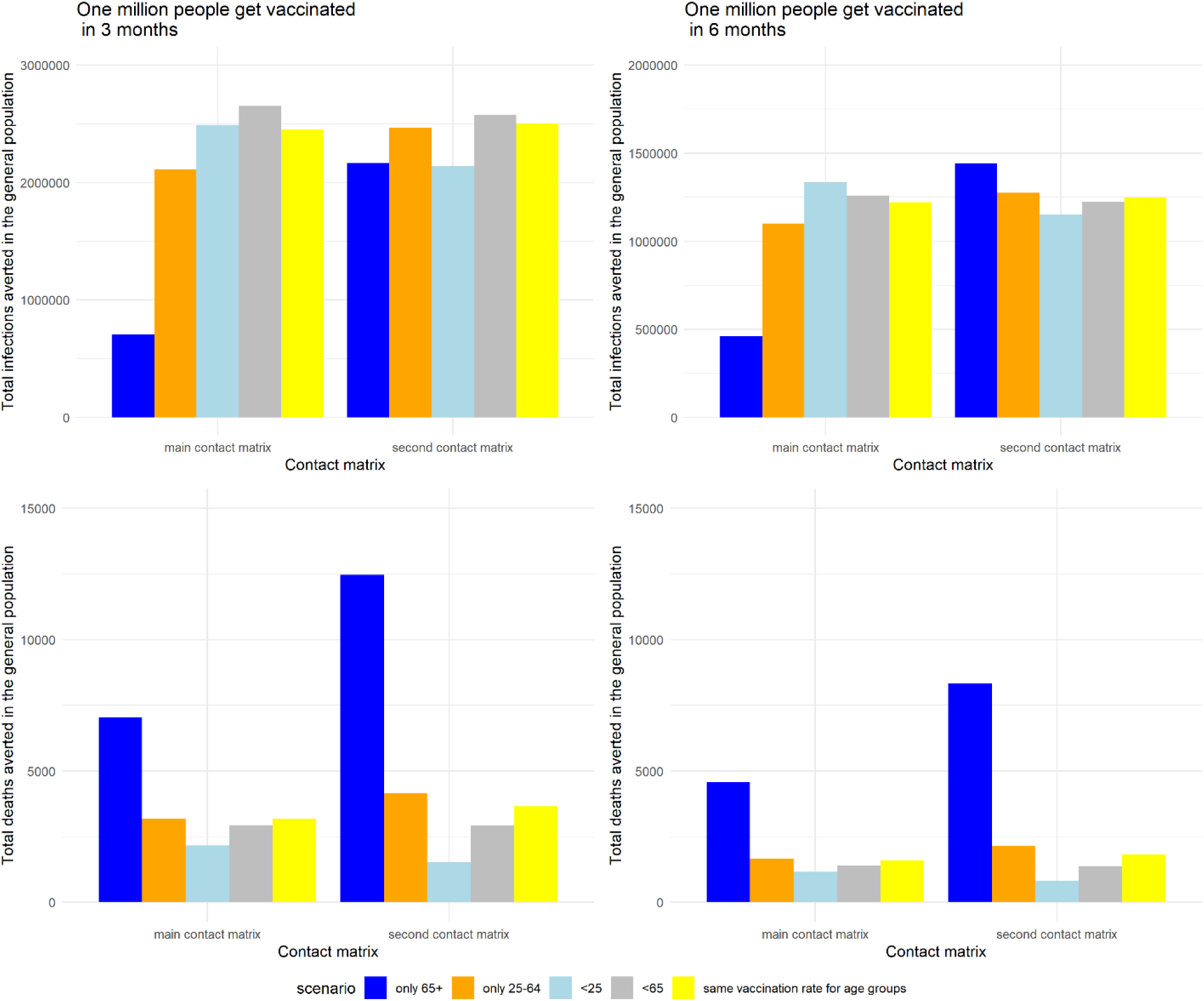
The impact of various vaccination scenarios on the number of cumulative infections averted (upper panel) and the number of deaths averted (lower panel) in the general population with two different contact matrices, in the main analysis with R0=3.13 (initial strain).

### Cumulative deaths averted under the main scenario of vaccination of one million persons

Vaccinating the elderly (65+ years) could avert over 7,000 deaths if one million people were vaccinated over three months, assuming the main contact matrix, while over 4,000 deaths would be averted if they were vaccinated over six months. The number of deaths prevented was the lowest when persons <25 years were prioritized in both vaccination time frames (2,171 in 3 months versus 1,157 in 6 months), assuming the main contact matrix (Figure 2). Vaccinating the elderly (65+ years) remained the best option to reduce deaths regardless of the mixing patterns (Figures 1 and 2; Tables 2 and 3).

### Varying vaccine supply to vaccinate 500,000 or two million persons

In contrast to the base case scenarios of vaccinating 1 million people, prioritizing persons <25 years was associated with the lowest percentage of cumulative infections if 500,000 or two million individuals were vaccinated in 3 or 6 months (Table 2). However, simulations using the second matrix reported mixed results. For example, prioritizing the elderly seemed to be the best strategy for lowering cumulative infections when vaccine supplies were only enough for 500,000 people (177.68% for three months and 179.54% for six months). In contrast, vaccinating each age group at the same rate was preferred when the supply was enough to vaccinate two million persons (166.10% for three months and 174.23% for six months) (Table 3). Prioritizing the elderly (65+ years) remained the strategy of choice to specifically lower COVID-19 mortality for both matrices (Tables 2 and 3).

### Comparing outcomes for the initial and delta variants in the absence of vaccination

In the absence of vaccination, the scenario analysis for the delta variant using the main matrix suggested the percentage of symptomatic persons at the peak, cumulative incidence, and deaths in the population at 10.29%, 231.24%, and 0.28%, respectively (Table S4), were higher than the initial strain at 7.26%, 174.37%, and 0.20%, respectively (Figure 1). This finding was consistent with the results from the second matrix (Table S5). In the delta variant scenario, the percentage of symptomatic individuals at the peak was slightly lower with the second matrix (10.14%) than with the main matrix (10.29%). However, the cumulative infections (238.73%) and deaths (0.31%) were higher with the second matrix in the delta variant scenario (Tables S4 and S5).

**Table 4:** Comparing outcomes for the delta variant when one million persons were vaccinated under the various vaccination strategies using the main matrix

| **Vaccine prioritization** | **Initial strain (3 months) (%)** | **Delta variant (3 months) (%)** | **Initial strain (6 months) (%)** | **Delta variant (6 months) (%)** |
| --- | --- | --- | --- | --- |
| **Symptomatic infections at peak** |  |  |  |  |
| Only 65+ | 7.19 | 10.22 | 7.22 | 10.25 |
| 25-64 | 6.92 | 10.08 | 7.09 | 10.18 |
| <25 | 6.79 | 10.01 | 7.01 | 10.14 |
| < 65 | 6.78 | 10.01 | 7.03 | 10.15 |
| Same vaccination rate across age groups | 6.83 | 10.03 | 7.04 | 10.16 |
| **Cumulative infections** |  |  |  |  |
| Only 65+ | 172.08 | 228.41 | 172.87 | 229.49 |
| 25-64 | 167.51 | 227.05 | 170.89 | 229.07 |
| <25 | 166.29 | 226.63 | 170.04 | 228.77 |
| < 65 | 165.76 | 226.25 | 170.28 | 228.87 |
| Same vaccination rate across age groups | 166.42 | 226.54 | 170.41 | 228.89 |
| **Deaths** |  |  |  |  |
| Only 65+ | 0.18 | 0.25 | 0.18 | 0.26 |
| 25-64 | 0.19 | 0.27 | 0.19 | 0.28 |
| <25 | 0.19 | 0.28 | 0.19 | 0.28 |
| < 65 | 0.19 | 0.27 | 0.19 | 0.28 |
| Same vaccination rate across age groups | 0.19 | 0.27 | 0.19 | 0.28 |

**Table 5:** Comparing outcomes for the delta variant when one million persons were vaccinated under the various vaccination strategies using the second matrix

| **Vaccine prioritization** | **Initial strain (3 months) (%)** | **Delta variant (3 months) (%)** | **Initial strain (6 months) (%)** | **Delta variant (6 months) (%)** |
| --- | --- | --- | --- | --- |
| **Symptomatic infections at peak** |  |  |  |  |
| Only 65+ | 6.92 | 9.96 | 7.02 | 10.04 |
| 25-64 | 6.76 | 9.90 | 6.96 | 10.01 |
| <25 | 6.82 | 9.91 | 6.98 | 10.01 |
| < 65 | 6.74 | 9.87 | 6.97 | 10.01 |
| Same vaccination rate across age groups | 6.76 | 9.89 | 6.98 | 10.01 |
| **Cumulative infections** |  |  |  |  |
| Only 65+ | 175.32 | 232.78 | 177.68 | 235.12 |
| 25-64 | 174.34 | 234.35 | 178.22 | 236.48 |
| <25 | 175.41 | 234.06 | 178.62 | 236.57 |
| < 65 | 174.00 | 232.32 | 178.39 | 236.50 |
| Same vaccination rate across age groups | 174.23 | 234.12 | 178.30 | 236.43 |
| **Deaths** |  |  |  |  |
| Only 65+ | 0.18 | 0.26 | 0.19 | 0.28 |
| 25-64 | 0.21 | 0.31 | 0.21 | 0.31 |
| <25 | 0.22 | 0.31 | 0.22 | 0.31 |
| < 65 | 0.21 | 0.31 | 0.22 | 0.31 |
| Same vaccination rate across age groups | 0.21 | 0.31 | 0.21 | 0.31 |

### Impact of vaccination strategies on symptomatic infections at the peak due to the delta variant

The vaccine prioritization findings for the delta variant scenario found that prioritizing persons <25 years was associated with the lowest percentage of symptomatic infections at the peak regardless of the available vaccine doses and rollout speed, using the main matrix (Table 4). Like the initial strain scenario, prioritizing persons <65 years was associated with the lowest percentage of symptomatic infections at the peak (9.87%) under the assumption of vaccinating one million persons over three months using the second matrix for the delta variant (Table 5).

### Impact of vaccination strategies on cumulative infections due to the delta variant

The scenario where one million people were vaccinated over three months suggested that focusing on persons <65 years had the lowest value of cumulative infections (226.25%) for the delta variant, like the initial strain (165.76%). Moreover, prioritizing persons below 25 years was the best option to minimize cumulative infections in the population with a 6-month rollout for the delta variant (228.77%) (Table 4). Importantly, the results on cumulative infections of the second matrix suggested the elderly (235.12%) should be prioritized for vaccination first for the delta variant with a 6-month rollout (Table 5).

### Impact of vaccination strategies on cumulative deaths due to the delta variant

Prioritizing the elderly remained the best strategy for lowering deaths in the population for the initial strain and the delta variant in all the scenarios (Tables 4, 5, S4, and S5).

## Discussion

Vaccination is the best tool to control the spread of SARS-CoV-2 and minimize the burden of COVID-19 globally. As Ghana relies primarily on multilateral donations for their COVID-19 vaccine supply, there is a need to determine the best vaccine optimization strategies to minimize deaths, cumulative case counts, or epidemic peaks over a relatively short period. Using two contact matrices, this study utilized an age-stratified mathematical model to answer the question “who should get vaccinated first?” when the vaccine supply was limited and when supplies were exhausted over three and six months. The findings suggest that, for both the initial strain and the delta variant, prioritizing persons <65 for vaccination would avert the most cumulative infections while prioritizing the elderly (65+ years) was associated with the lowest death counts.

Optimization of vaccine prioritization strategy is sensitive to the population structure. The finding of prioritizing younger persons to avert cumulative infections has been reported in other studies (*18, 33, 34*). Bubar and colleagues concluded in their multi-country research that the cumulative incidence of COVID-19 was lowest when adults between 20-49 years were prioritized, especially with a highly-effective transmission-blocking vaccine (*18*). In Senegal, Diarra and colleagues used an age-structured dynamic mathematical model to explore various vaccination strategies and reported that prioritizing persons 60 years and below was associated with the lowest case burden (*35*). The authors argued that countries with younger populations, like Ghana, should prioritize vaccinating younger persons to minimize hospital costs and productivity loss.

Similar to our findings, these studies also concluded that prioritizing the elderly was associated with the lowest mortality. Regardless, Bubar and colleagues reported that persons between 20-49 years should be prioritized to minimize mortality when the transmission was low, vaccine efficacy was lower in older adults, and when the vaccine had a high effect in blocking transmission. Buckner and colleagues also reported similar results to our study and found that to control deaths directly, the elderly should be vaccinated first after stratifying young adults by essential worker status (*36*). Although the conclusions in both studies were similar, Buckner and colleagues used a dynamic approach in modeling vaccine allocation strategies that accounted for changes in the epidemiologic status of the population (“shares of the population in different disease states”) over six months using stochastic nonlinear programming techniques. Similar to our findings, in their vaccine optimization modeling study in India, Foy and colleagues concluded that prioritizing older adults (i.e., 60+ years) was associated with the most significant reduction in deaths regardless of vaccine efficacy, control measures, and rollout speed (*33*). Another modeling study by Chapman and colleagues using COVID-19 data from California reported similar results (*37*). However, in the Chapman study, they focused on identifying the groups to prioritize after healthcare workers and long-term facility residents received initial vaccine doses.

The differences in outcomes between the contact matrices may be due to the significantly lower reported contact rates among the younger population in the matrix adapted from Trentini et al. (i.e., “second matrix”) (*29*). The study by Bubar and colleagues on vaccine optimization strategies across multiple countries, including South Africa, concluded the best vaccination strategy depended on the extent of mixing patterns (*18*). Future studies may consider exploring differences observed using matrices of different settings, i.e., rural versus urban and household versus community mixing.

As reported by Ko and colleagues, the question of whom to be vaccinated first also depended on the objective for vaccination (minimizing cumulative infections or deaths) and the effective reproduction number (*34*). Thus, policymakers may need to consider the compromises in deciding the best vaccine allocation strategies. For example, prioritizing the elderly may lead to fewer deaths but higher case numbers, which could exacerbate economic loss due to a high case count in a younger population. Moreover, the transmissibility of the circulating variant may also inform the vaccine optimization strategy. However, this was not evident in our study as the priority group remained the same for the delta variant with a higher reproduction number. As found in some of our scenario analyses, other studies concluded vaccine optimization also depended on the vaccine supply (*37, 38*).

Although this study has demonstrated the need to prioritize certain age groups to minimize the burden of COVID-19 in Ghana depending on the objective of the program, other factors need to be considered to ensure people receive vaccinations when they become eligible. It might be imperative to adopt targeted vaccine campaigns to minimize hesitancy in the group to be prioritized. For example, Acheampong and colleagues reported the level of reluctance among older adults was lower compared to younger adults in Ghana (*39*). Likewise, a survey among persons >65 years in the US also found that 91% of the elderly were willing to get vaccinated (*40*). Given the reported vaccine hesitancy by age group, there is a need for campaigns to create an enabling environment and engage the younger population about their role in mitigating the pandemic.

Our study has several limitations. First, our model was age-stratified only. Other demographic variables (e.g., occupation and comorbidity) may change one’s COVID-19 infection risk and clinical prognosis (*41*). Second, the model did not include a hospitalization compartment. Thus, the impact of the vaccine in the context of the omicron variant with demonstrated effectiveness against severe infections but modest effectiveness against infection could not be studied. Third, the contact matrices used were adapted from other African countries. These countries had similar demographic distributions as Ghana, and we assumed that the frequency of contact in the population would be similar. Fourth, we did not have data to represent the rural and urban differences in contact matrices in Ghana. Fifth, our model design specified the symptomatic period was the same for individuals who recovered from COVID-19 and those who died of it. Sixth, the highly transmissible omicron variant was not included in our study due to limited evidence of the effectiveness of vaccines against infection from the variant (*42*). Lastly, our model accounted for vaccine effectiveness against infection but did not account for the reduction of the case-fatality ratio if one was vaccinated yet infected.

In conclusion, we used an age-stratified compartmental model to assess the impact of various COVID-19 vaccine allocation strategies in Ghana. This study reiterates the need to increase vaccination rates by ensuring increased vaccine supplies and faster rollout speed. Vaccinating persons <65 or <25 years was associated with the highest numbers of cumulative infections averted for the initial strain and the delta variant while prioritizing persons 65+ years was associated with the lowest deaths in the population. The findings indicated that vaccine prioritization strategies in a country are dependent on its policy objective, population structure, mixing patterns, and vaccine supply.

## First author bio

Dr. Sylvia K. Ofori graduated from Georgia Southern University with her Doctor of Public Health degree with a concentration in epidemiology. Her research interests are infectious disease epidemiology and mathematical modeling.

## Author contributions

S.K.O. and I.C.H.F. conceptualized the study; S.K.O. programmed the model in R and ran the simulations and drafted the manuscript under I.C.H.F.’s supervision. J.S.S., K.L.S., G.C., and B.J.C. provided consultation to S.K.O. and I.C.H.F. All authors edited, reviewed, and approved the manuscript.

## Conflict of interest statement

Sylvia Ofori declares that she is a paid intern at Ionis Pharmaceuticals, and the financial relationship does not affect the content of the article. Prof. Benjamin Cowling declares that he was a consultant for Roche and Sanofi Pasteur. All other co-authors declare no competing interest.

## Funding statement

This project is not supported by any external funding agencies.

## Data Availability

This is an epidemiological modeling study. We have provided the source R code in Technical Appendix B so that others can reproduce our results.

## Technical Appendix A

### Supplementary Text: Methods

#### Model equations

The total population of Ghana, *N*, is assumed to be a constant and equals the sum of the compartments representing individuals with different disease statuses (including COVID-19- specific deaths) at any time during the simulation and that births and deaths from natural (non-COVID-19) causes do not affect the infection dynamics in the population:

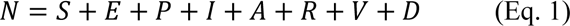

Following the assumption of a frequency-dependent model, we define the force of infection, *λ*, as a combination of infection terms with symptomatic, asymptomatic, and pre-symptomatic individuals, each contributing to the transmission process as follows:

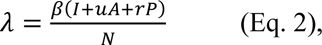

where the contributions of asymptomatic and presymptomatic individuals are fractions *u* and *r* of that of the symptomatic individuals.

The set of ordinary differential equations (ODE) that defines the progression of susceptible individuals through different disease statuses upon infection and a vaccinated and immune status upon vaccination and their re-entry into the susceptible state due to waning immunity is described below:

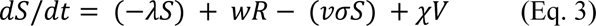

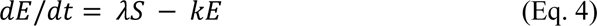

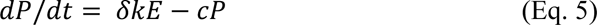

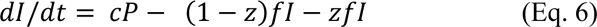

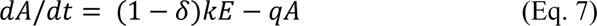

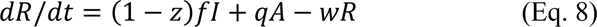

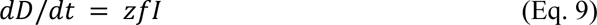

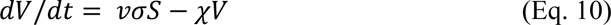

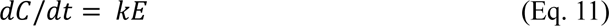

In the SEPIARD-V model, the population is initially susceptible until an infectious individual is introduced. After contact with an infectious person, susceptible individuals are infected at a rate of *λ* (force of infection). While in the latent period (E), they do not transmit the virus. Individuals leave the latent period at a rate of *k* and can either become asymptomatically (A) or pre-symptomatically infectious (P). Asymptomatic individuals will recover (move to the R compartment) at a rate of *q* without showing any symptoms (*1, 2*). Pre-symptomatic infectious individuals become symptomatic (I) at a rate of *c*. The mean duration of the symptomatic period is defined as 1/*f*. A fraction *z* of symptomatic individuals will die from COVID-19 (move to the D compartment) while the other fraction (1-*z*) will recover (move to the R compartment). Susceptible individuals become fully vaccinated (move to the V compartment) at a rate of *v* per day, while the vaccine is assumed to have an efficacy (or effectiveness) of *σ*.

#### Model parameters

In our model, after susceptible individuals are exposed, the latent period, which is the period from exposure to infectiousness, is 1/*k* and is assumed to have a mean of 1.85 days (*3, 4*). Once exposed, a third (*δ*) of individuals become pre-symptomatically infected, and the rest (1-δ) become asymptomatic (*5, 6*). The mean pre-symptomatic period, 1/*c*, is assumed to be 2.9 (*7*). The mean duration of infectiousness for symptomatic individuals (1/*f*) is 15.7 days, and that of asymptomatic individuals (1/*q*) is 7.25 days (*8-11*). The transmission rate, *β*, is estimated from the basic reproduction number (R_0_) using the formula 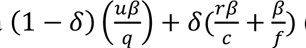(*12*), assuming a value of 3.13 for the initial strain as assessed by Armachie and colleagues (*13*). This value would be updated in our scenario analysis of the delta variant. According to the CDC COVID-19 pandemic planning scenarios, the relative transmissibility of asymptomatic and pre-symptomatic individuals, *u* and *r*, are assumed to be 0.75, respectively (*14*). Two doses of the AstraZeneca COVID-19 vaccine were reported to have an efficacy (*σ*) of 0.745 (*15*). This value would also be updated in our scenario analysis of the delta variant. Immunity is acquired from either natural infection or vaccination. Vaccination-induced immunity offers protection from infection for six months (180 days) and wanes at a rate of *χ*, while that from natural infection, *w*, is about one year (365 days) (*16*). The rate of vaccination, *v*, is varied depending on the scenario. Once immunity wanes, individuals move back to the susceptible compartment. Details of model parameters are found in Table 1.

#### Age-stratification

The aforementioned SEPIARD-V model was further developed into an age-stratified model. The idea of the age-stratified model was adapted from a modeling study by Keeling and White on vaccination strategies with an optimal number of cases and severity effects during Britain’s 2009 H1N1 influenza pandemic (*17*). Our analysis would answer research questions similar to the Keeling and White study and include modifications to address issues pertinent to the COVID-19 pandemic in Ghana. With vaccine supplies available, policymakers would be interested in which epidemiological goal the vaccine would most impact.

A recent retrospective cohort study in Ghana by Ashninyo et al. reported that COVID-19 disproportionately affected the younger population with a mean age of 37.9 years, with the majority (56.64%) between 31 and 64 years (*18*). According to Ghana’s demographics, 56.08 % of the population is below 25 years, and 4.44% are 65 years or above (*19*). Therefore, the population was stratified into three groups: <25 years, 25-64 years, and 65+ years.

#### Age-stratified model formulation

An age-stratified compartmental model assumes that population mixing is not homogeneous and the numbers of contact between members of age groups follow a specified contact matrix. The number of secondary cases caused by an infectious individual in a totally susceptible population is commonly known as the basic reproduction number. In the context of a heterogeneous-mixing model, the basic reproduction number is also known as a basic reproductive ratio and is the largest eigenvalue of the next generation matrix (NGM) (*20*). Following the work of Towers and Feng (*21*), the reproduction number of an age-stratified model is equal to the product of the transmission coefficient β, the mean duration of infectiousness, and the largest eigenvalue of a matrix M that is defined by its elements 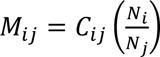, where C_ij_ is the contact matrix, and N_i_ and N_j_ are the numbers of individuals in age groups i and j respectively (*22*).

#### Contact matrices used

Due to the strong evidence of assortative mixing between age groups in the general population of Uganda (*23*) and Kenya (*24*), the contact matrix of the population was considered in the modeling of vaccination allocation strategies in Ghana. As reported by Waroux and colleagues, the contact patterns of Uganda were adopted in this study because their matrix corresponds to the population groups used in this study (below 25 years, 25-64 years, and 65 years or above). There is also a similarity in the proportion of age structure between Uganda and Ghana. Waroux and colleagues used the survey method to study the contact patterns of residents in rural Uganda in 2014 and found that, on average, the within-group contact rate among individuals below 25 years is 23.58 per day; for those between 25-64 years, it was 15.05 per day and 0.54 per day for those above 64 years (*23*).

Therefore, the 3 by 3 contact matrix is:

|  | **Below 25 years** | **25-64 years** | **65 years and above** |
| --- | --- | --- | --- |
| **Below 25 years** | 23.58 | 9.31 | 0.87 |
| **25-64 years** | 13.01 | 15.05 | 1.53 |
| **65 years and above** | 2.29 | 2.44 | 0.54 |

This contact matrix was corrected for reciprocity using methods described by Melegaro et al. in their study in Zimbabwe (*25*).

The second matrix was adapted from a study in Ethiopia by Trentini et al., who also used survey-type interviews to estimate age-specific patterns (*26*). The contact matrix was used due to the similar population structure to Ghana. Furthermore, the data on contact patterns were collected in 2019, prior to the COVID-19 pandemic, and may reflect recent contact rates. On average, the within-group contact rate among individuals below 25 years is 8.2 per day; for those between 25- 64 years, it was 7.8 per day, and 1.6 per day for those above 64 years (*23*). The 3 by 3 contact matrix is:

|  | **Below 25 years** | **25-64 years** | **65 years and above** |
| --- | --- | --- | --- |
| **Below 25 years** | 8.2 | 5 | 1 |
| **25-64 years** | 2 | 7.8 | 2.8 |
| **65 years and above** | 0.1 | 2.2 | 1.6 |

This contact matrix was then corrected for reciprocity using methods described by Melegaro in their study in Zimbabwe (*27*).

#### Case-fatality ratio in the age-stratified model

The age-specific fatality ratios were calculated using data from Odikro and colleagues’ study on the epidemiology of COVID-19 outbreak in Ghana (*28*). Using the total number of cases reported in their study (n=17,763) and the percentage of cases reported in each 10-year age group as of June 30, 2020, we calculated the percentage of cases in each age group as 23.85% for persons below 25 years, 70.65% for those between 25-64 years and 5.5% for 65+ years. For the cases reported among persons 20-29 years, we assumed that half of them occurred in persons between 20-24 years, and the other half occurred in those between 25-29 years. Next we calculated the expected number of cases for <25 years (n=4,236), 25-64 years (n=12,550) and elderly (n=977). The expected number of deaths was estimated for each age group assuming that 9% of the total deaths (n=117) deaths occurred among <25 years, 51% for 25-64 years and, 40% among the elderly (65+) (*28, 29*). Finally, we calculated the age-specific case fatality ratios as the ratio between the number of deaths in each age group by the number of cases in each age group. Hence, the estimates were 0.002 for <25 years, 0.005 for 25-64 years and 0.048 for 65+ years. All other variables except the vaccination rate remained the same as described in Table 1.

#### Model initialization

The model’s system of ODE was solved following the Runge-Kutta 4 method in the deSolve package in R version 4.1.1 (*30*). To keep it simple, the population size of Ghana, N, was set to 30,800,000. We also assumed that for the base case scenario, at the beginning of the simulation, I =1, A=0, P=0, D=0, and V=0. We accounted for the age-specific seroprevalence of SARS-CoV-2 using estimates from Quarshie and colleagues in August 2020 (*31*). We, therefore, assumed that 17.5% of persons below 25 years, 43.6% of those between 25-64 years, and 18% of 65+ persons had been infected at the beginning of the simulation. These individuals were moved to the recovery compartment at the beginning of the simulation. The model was run for 500 days to allow enough time for the first wave of the epidemic to die out and observe when the second wave began to emerge.

#### Outcomes

The cumulative number of infections and deaths averted in the general population was estimated and compared for each scenario. Furthermore, the percent of the population who were symptomatic at the peak, ever infected (cumulative infections), and cumulative deaths were assessed. The percentage of cumulative infection could exceed 100% because as immunity waned, individuals would become susceptible again to repeated infections.

#### R code

The R code used for simulation in this study is provided in Technical Appendix B.

Daily vaccination rates for vaccinating one million people in three months and six months using an age-stratified model

**Table S1: Daily vaccination rates for vaccinating one million people in three months and six months using an age-stratified model**
| **Scenario** | **Fraction of total population** | **Number in each subgroup** | **1 million people can be vaccinated, % of the subpopulation vaccinated** | **The daily vaccination rate for a campaign of 3 months** | **The daily vaccination rate for a campaign of 6 months** |
| --- | --- | --- | --- | --- | --- |
| Only 65+ y | 0.044 | 1,355,200 | 73.8% | 0.008199 | 0.00410 |
| 25-64 y | 0.396 | 12,196,800 | 8.2% | 0.000911 | 0.00046 |
| <25 y | 0.56 | 17,248,000 | 5.8% | 0.000644 | 0.00032 |
| <65 y | 0.956 | 29,444,800 | 3.4% | 0.000377 | 0.00019 |
| Same vaccination rate | 1 | 30,800,000 | 3.2% | 0.000361 | 0.00018 |

Daily vaccination rates for vaccinating 500,000 people in three months and six months using an age-stratified model

**Table S2: Daily vaccination rates for vaccinating 500,000 people in three months and six months using an age-stratified model**
| **Scenario** | **Fraction of total population** | **Number in each subgroup** | **500,000 people can be vaccinated, % of the subpopulation vaccinated** | **The daily vaccination rate for a campaign of 3 months** | **The daily vaccination rate for a campaign of 6 month** |
| --- | --- | --- | --- | --- | --- |
| Only 65+ y | 0.044 | 1,355,200 | 36.9% | 0.004099436 | 0.00205 |
| 25-64 y | 0.396 | 12,196,800 | 4.1% | 0.000455493 | 0.00023 |
| <25 y | 0.56 | 17,248,000 | 2.9% | 0.000322099 | 0.00016 |
| <65 y | 0.956 | 29,444,800 | 1.7% | 0.000188677 | 0.00009 |
| Same vaccination rate | 1 | 30,800,000 | 1.6% | 0.000180375 | 0.00009 |

**Table S3:** Daily vaccination rates for vaccinating two million people in three months and six months using an age-stratified model

| **Scenario** | **Fraction of total population** | **Number in each subgroup** | **2 million people can be vaccinated, % of the subpopulation vaccinated** | **The daily vaccination rate for a campaign of 3 months** | **The daily vaccination rate for a campaign of 6 months** |
| --- | --- | --- | --- | --- | --- |
| Only 65+ y | 0.044 | 1355200 | 147.6% | 0.0163977 | 0.00819887 |
| 25-64 y | 0.396 | 12196800 | 16.4% | 0.001822 | 0.00091099 |
| <25 y | 0.56 | 17248000 | 11.6% | 0.0012884 | 0.0006442 |
| <65 y | 0.956 | 29444800 | 6.8% | 0.0007547 | 0.00037735 |
| Same vaccination rate | 1 | 30800000 | 6.5% | 0.0007215 | 0.00036075 |

**Table S4:**
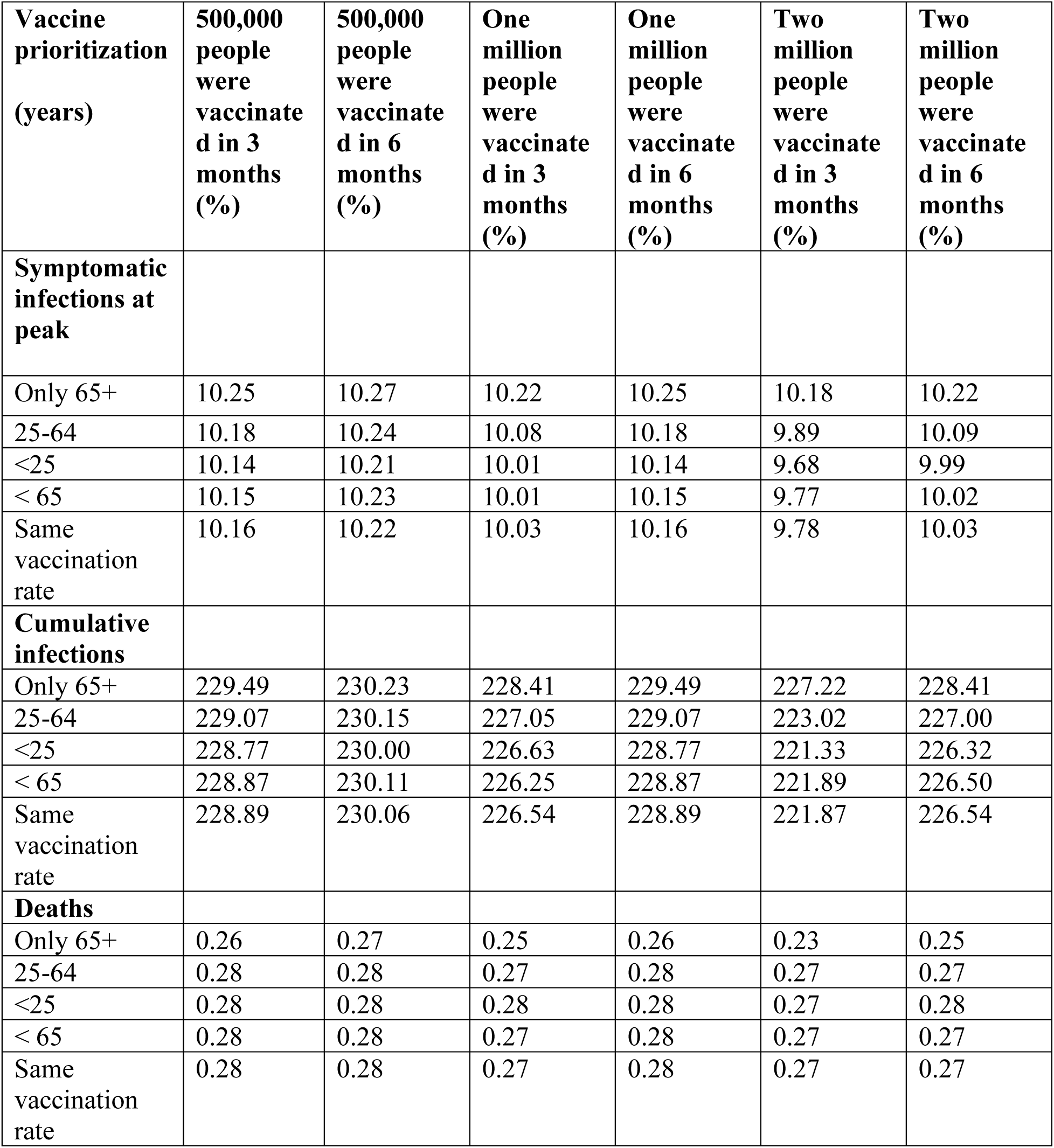
Sensitivity analysis of outcomes in the total population under various vaccination scenarios using the main matrix for the delta variant. If there were no vaccination, 10.29% of the population would be symptomatic at the epidemic peak, there would be a total of 231.24% cumulative incidence, and 0.28% of the population would die of COVID-19.

| **Vaccine prioritization (years)** | **500,000 people were vaccinated in 3 months (%)** | **500,000 people were vaccinated in 6 months (%)** | **One million people were vaccinated in 3 months (%)** | **One million people were vaccinated in 6 months (%)** | **Two million people were vaccinated in 3 months (%)** | **Two million people were vaccinated in 6 months (%)** |
| --- | --- | --- | --- | --- | --- | --- |
| **Symptomatic infections at peak** |  |  |  |  |  |  |
| Only 65+ | 10.25 | 10.27 | 10.22 | 10.25 | 10.18 | 10.22 |
| 25-64 | 10.18 | 10.24 | 10.08 | 10.18 | 9.89 | 10.09 |
| <25 | 10.14 | 10.21 | 10.01 | 10.14 | 9.68 | 9.99 |
| < 65 | 10.15 | 10.23 | 10.01 | 10.15 | 9.77 | 10.02 |
| Same vaccination rate | 10.16 | 10.22 | 10.03 | 10.16 | 9.78 | 10.03 |
| **Cumulative infections** |  |  |  |  |  |  |
| Only 65+ | 229.49 | 230.23 | 228.41 | 229.49 | 227.22 | 228.41 |
| 25-64 | 229.07 | 230.15 | 227.05 | 229.07 | 223.02 | 227.00 |
| <25 | 228.77 | 230.00 | 226.63 | 228.77 | 221.33 | 226.32 |
| < 65 | 228.87 | 230.11 | 226.25 | 228.87 | 221.89 | 226.50 |
| Same vaccination rate | 228.89 | 230.06 | 226.54 | 228.89 | 221.87 | 226.54 |
| **Deaths** |  |  |  |  |  |  |
| Only 65+ | 0.26 | 0.27 | 0.25 | 0.26 | 0.23 | 0.25 |
| 25-64 | 0.28 | 0.28 | 0.27 | 0.28 | 0.27 | 0.27 |
| <25 | 0.28 | 0.28 | 0.28 | 0.28 | 0.27 | 0.28 |
| < 65 | 0.28 | 0.28 | 0.27 | 0.28 | 0.27 | 0.27 |
| Same vaccination rate | 0.28 | 0.28 | 0.27 | 0.28 | 0.27 | 0.27 |

**Table S5:** Sensitivity analysis of outcomes in the total population under various vaccination scenarios using the second matrix for the delta variant. If there were no vaccination, 10.14% of the population would be symptomatic at the epidemic peak, there would be a total of 238.73% cumulative incidence, and 0.31% of the population would die of COVID-19.

| **Vaccine prioritization** | **500,000 people were vaccinated in 3 months (%)** | **500,000 people were vaccinated in 6 months (%)** | **One million people were vaccinated in 3 months (%)** | **One million people were vaccinated in 6 months (%)** | **Two million people were vaccinated in 3 months (%)** | **Two million people were vaccinated in 6 months (%)** |
| --- | --- | --- | --- | --- | --- | --- |
| **Symptomatic infections at peak** |  |  |  |  |  |  |
| Only 65+ | 10.04 | 10.09 | 9.96 | 10.04 | 9.85 | 9.96 |
| 25-64 | 10.01 | 10.08 | 9.90 | 10.01 | 9.66 | 9.89 |
| <25 | 10.01 | 10.08 | 9.91 | 10.01 | 9.64 | 9.89 |
| < 65 | 10.01 | 10.08 | 9.87 | 10.01 | 9.64 | 9.89 |
| Same vaccination rate | 10.01 | 10.08 | 9.89 | 10.01 | 9.64 | 9.89 |
| **Cumulative infections** |  |  |  |  |  |  |
| Only 65+ | 235.12 | 236.68 | 232.78 | 235.12 | 230.20 | 232.78 |
| 25-64 | 236.47 | 237.60 | 234.35 | 236.48 | 230.09 | 234.30 |
| <25 | 236.57 | 237.65 | 234.71 | 236.57 | 230.11 | 234.44 |
| < 65 | 236.51 | 237.67 | 234.06 | 236.51 | 229.99 | 234.29 |
| Same vaccination rate | 236.43 | 237.58 | 234.12 | 236.43 | 229.52 | 234.12 |
| **Deaths** |  |  |  |  |  |  |
| Only 65+ | 0.28 | 0.30 | 0.26 | 0.28 | 0.24 | 0.26 |
| 25-64 | 0.31 | 0.31 | 0.31 | 0.31 | 0.30 | 0.31 |
| <25 | 0.31 | 0.31 | 0.31 | 0.31 | 0.31 | 0.31 |
| < 65 | 0.31 | 0.31 | 0.31 | 0.31 | 0.30 | 0.31 |
| Same vaccination rate | 0.31 | 0.31 | 0.31 | 0.31 | 0.30 | 0.31 |

**Figure S1:**
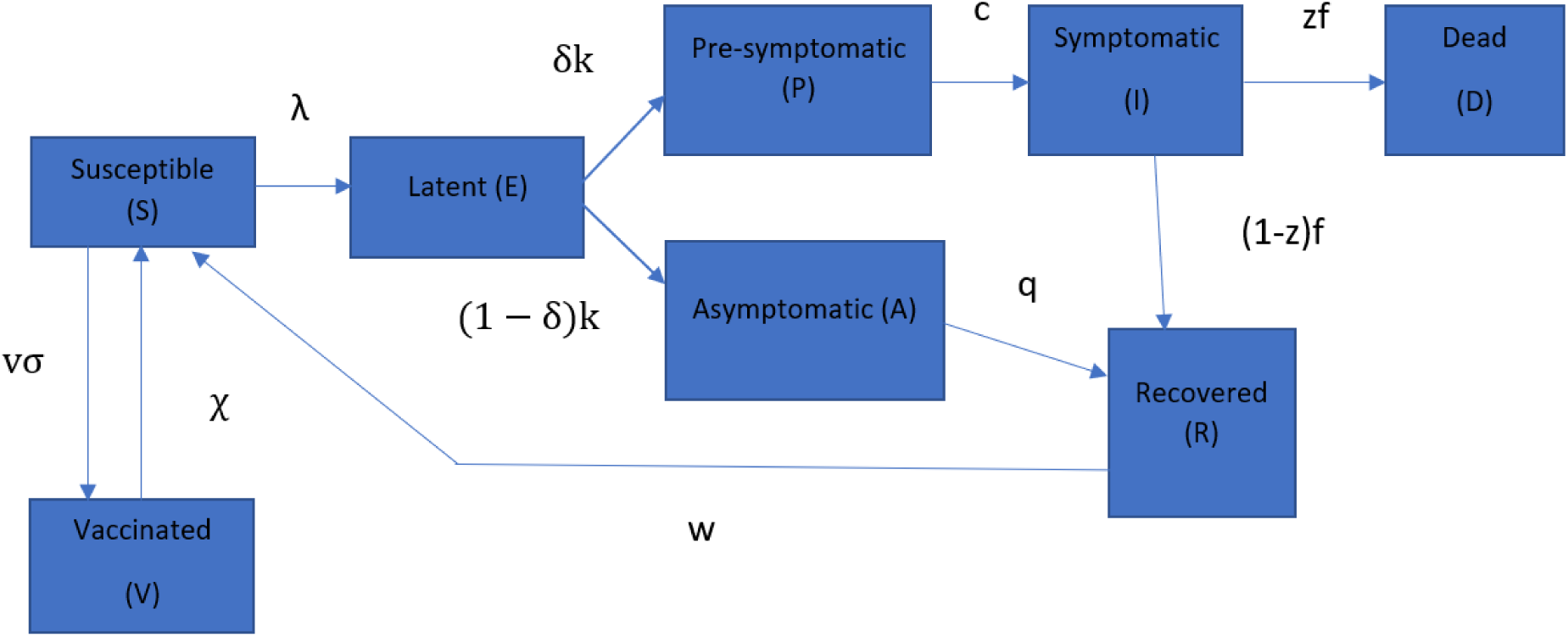
The Susceptible-Exposed-Presymptomatic-Symptomatic-Asymptomatic-Recovered-Dead-Vaccinated (SEPIARD-V) model represents SARS-CoV-2 transmission and COVID-19 disease progress and the vaccination against COVID-19.

## Technical Appendix B

First we clear the workspace to get rid of leftover variables.

`rm(list=ls())`

Next we clear all graphic windows.

`graphics.off()`;

We load packages that are required for the R simulation.

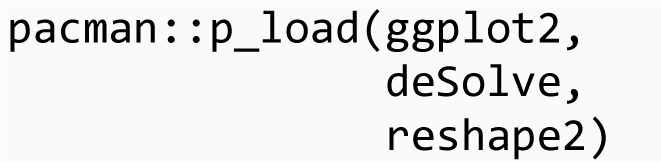

We set a seed so that the same simulation outputs can be obtained when simulations are repeated.

`set.seed(1234)`

### SEAPIRD-V model

- S refers to susceptible individuals.
- E refers to exposed individuals who are in the latent state.
- A refers to asymptomatic individuals: A percentage of the infected population makes transitions into the asymptomatic state. They are infectious.
- P refers to pre-symptomatic individuals: A percentage of the infected population makes transitions into the pre-symptomatic state. They are infectious.
- I refers to symptomatic individuals. They are infectious.
- R refers to recovered: Asymptomatic and Symptomatic individuals enter the recovery state.
- D refers to symptomatic individuals who died from COVID-19.
- V refers to individuals who become immune from COVID-19 infection after receiving two doses of the AstraZeneca vaccine.

### Parameter definitions

Parameters:

- c: transition rate from presymptomatic to symptomatic= 1/(incubation period - latency period)
- beta: transmission coefficient
- q: recovery rate for asymptomatic individuals
- delta: probability that exposed persons become presymptomatically infected
- f: the inverse of the duration of individuals being symptomatic (equals to the recovery rate for symptomatic individuals)
- u: relative transmissibility of asymptomatic individuals
- r: relative transmissibility of pre-symptomatic individuals
- sigma: vaccine efficacy
- v: vaccination rate
- chi: waning immunity of vaccinated individuals
- Ro: reproduction number
- z: age-specific case fatality ratio

In this study, two strains of COVID-19 were assessed by varying the reproduction numbers and vaccine efficacy with the values below

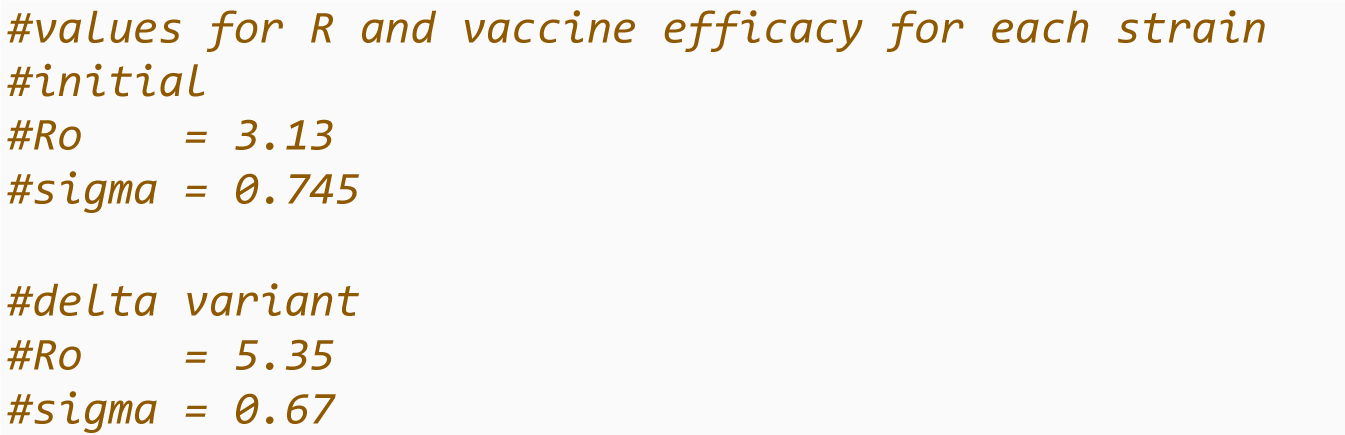

An age-specific case fatality ratio was calculated: We assumed 9% of deaths in the population occurred in children, 51% between 25 and 64 years, and 40% of deaths in the elderly. These translated into the following CFR: <25 yr = 0.002; 25-64 yr = 0.005; 65+ yr = 0.048. See Technical Appendix A for more details.

`z= c(0.002, 0.005, 0.048)`

We created a function for the age-stratified model with 9 compartments with the set of ordinary differential equations that defines the model.

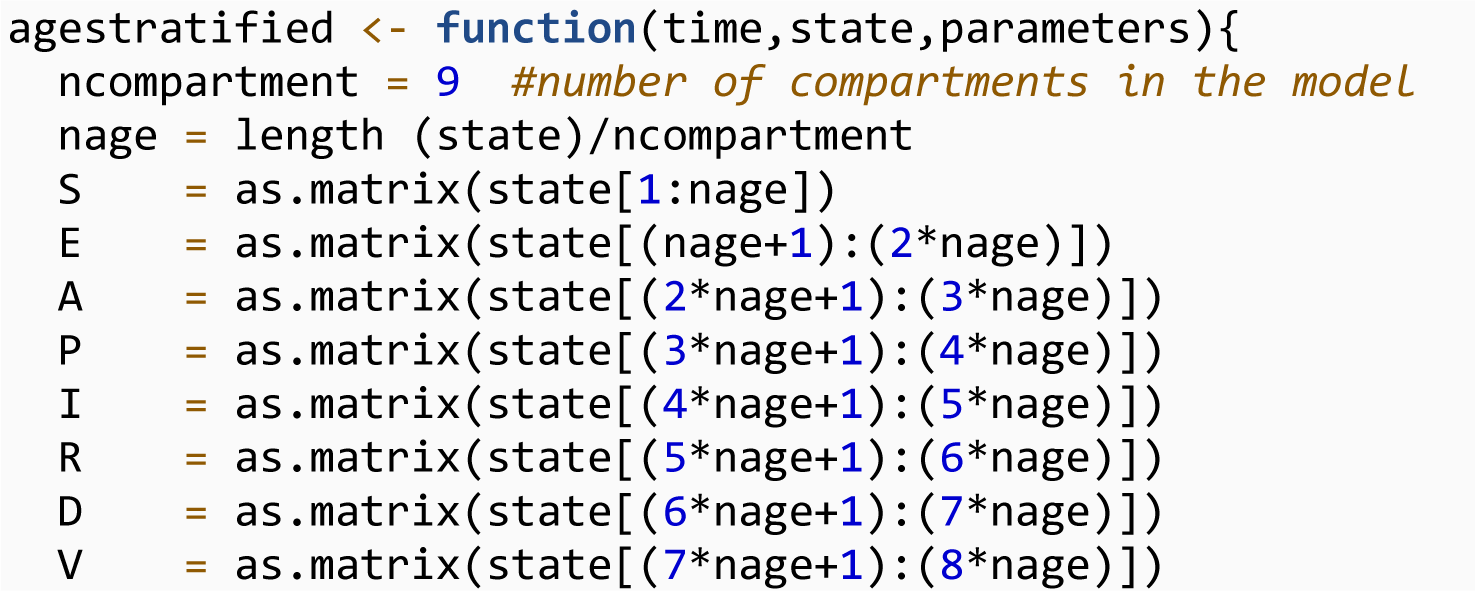
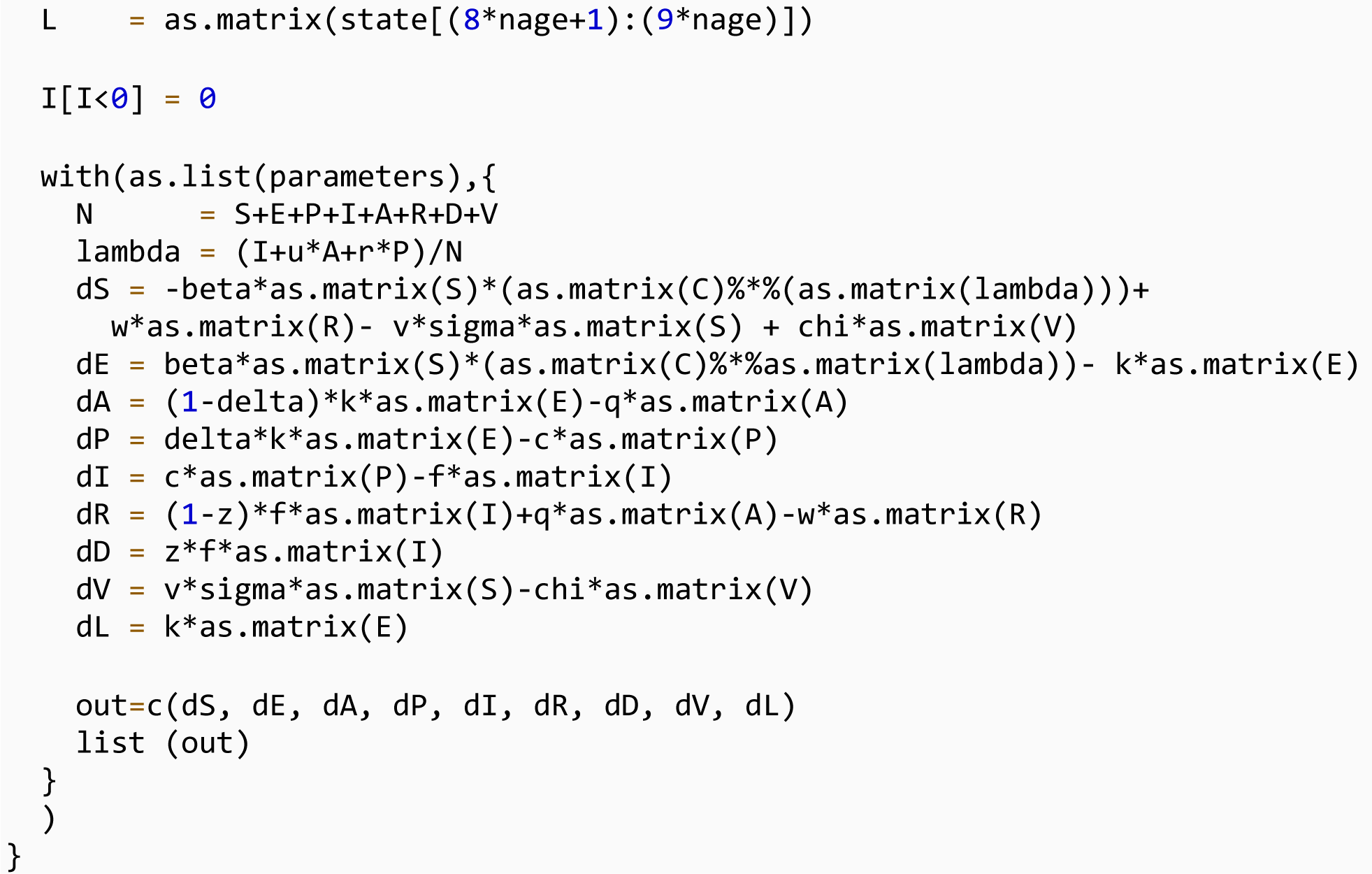

We determined the number of persons in each age group assuming a population size of 30800000.

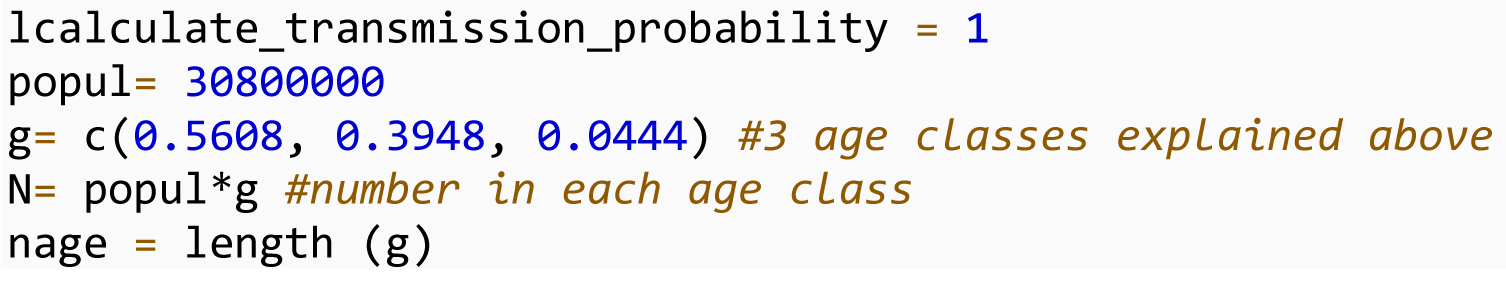

### Setting initial state for compartments

We assumed a % of each subpopulation was already infected, hence they were in the recovered compartment at the beginning of the simulation.

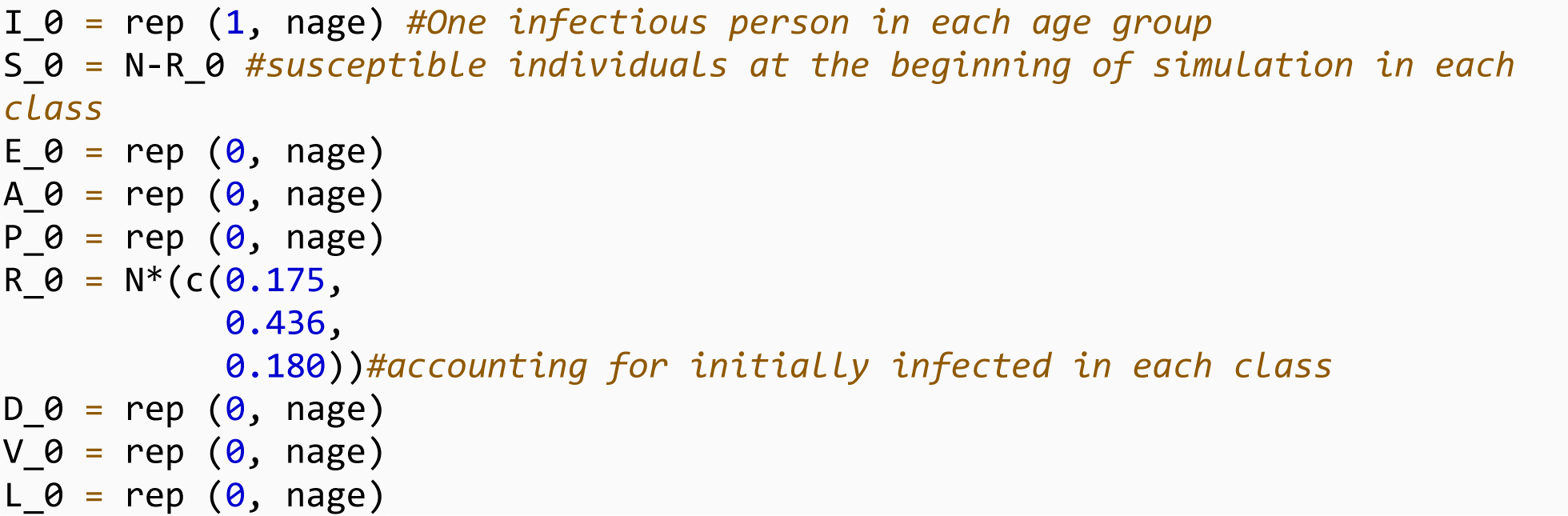

Two contact matrices were used in the study a) the first matrix is defined as the “main matrix”.

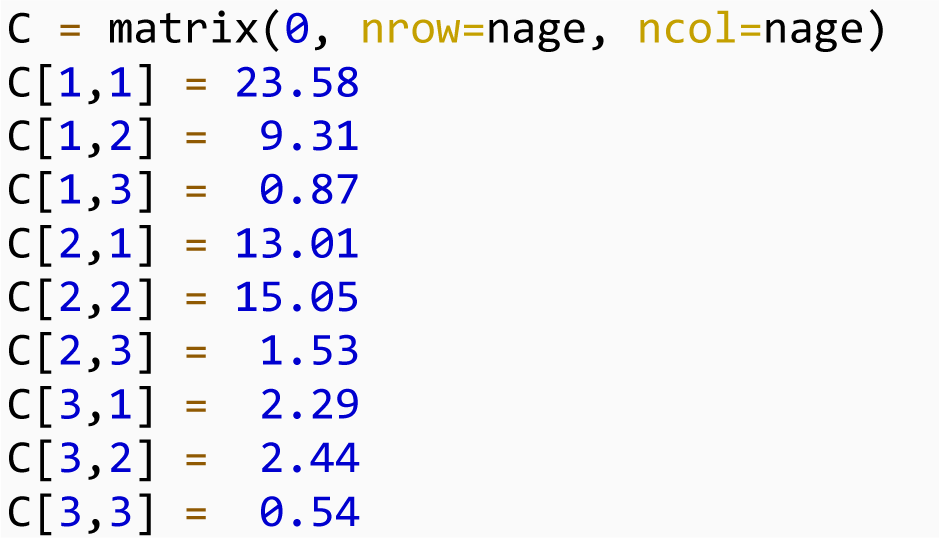

b) The other matrix is defined as “second matrix”

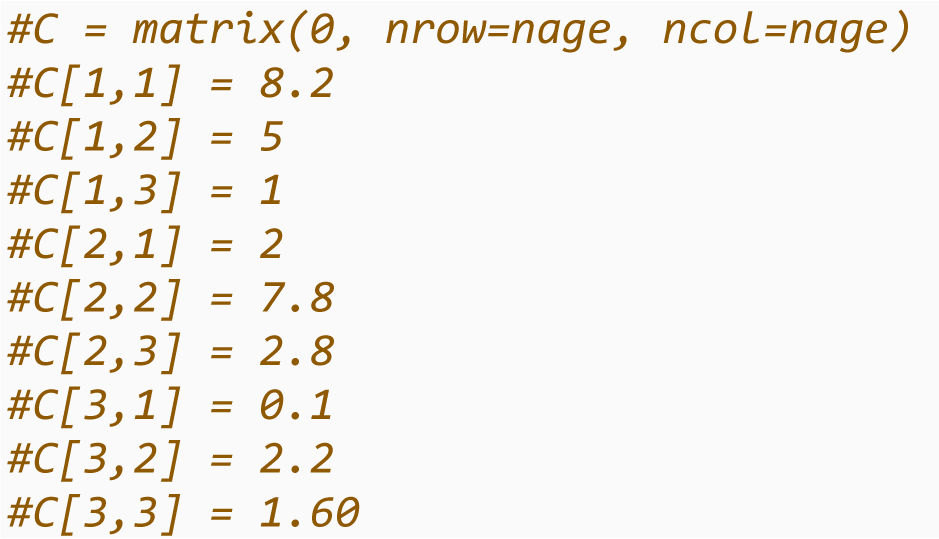

We set the parameter values used for calculating beta.

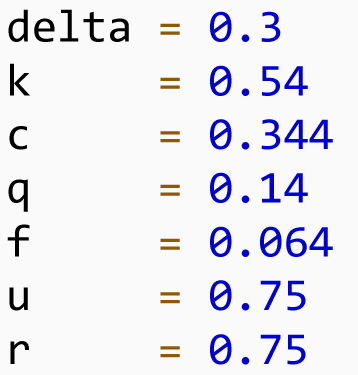

We defined the formulae for calculating the beta using the R0 from the largest eigenvalue.

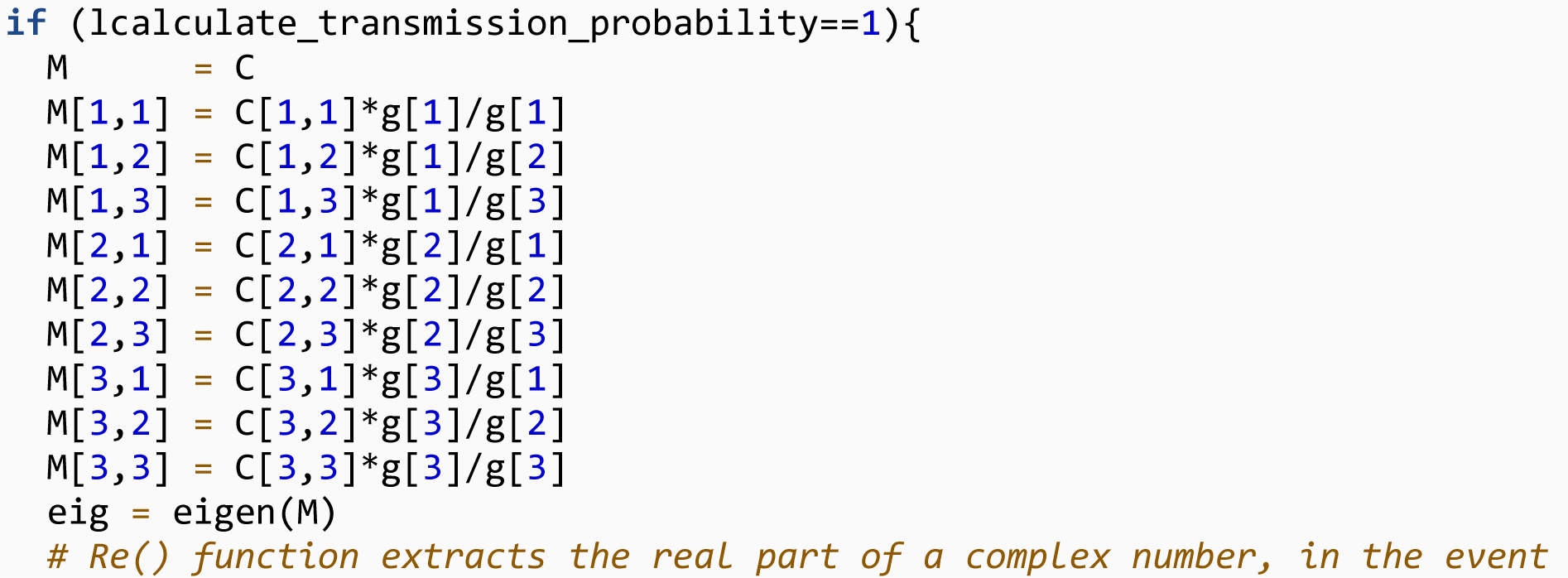
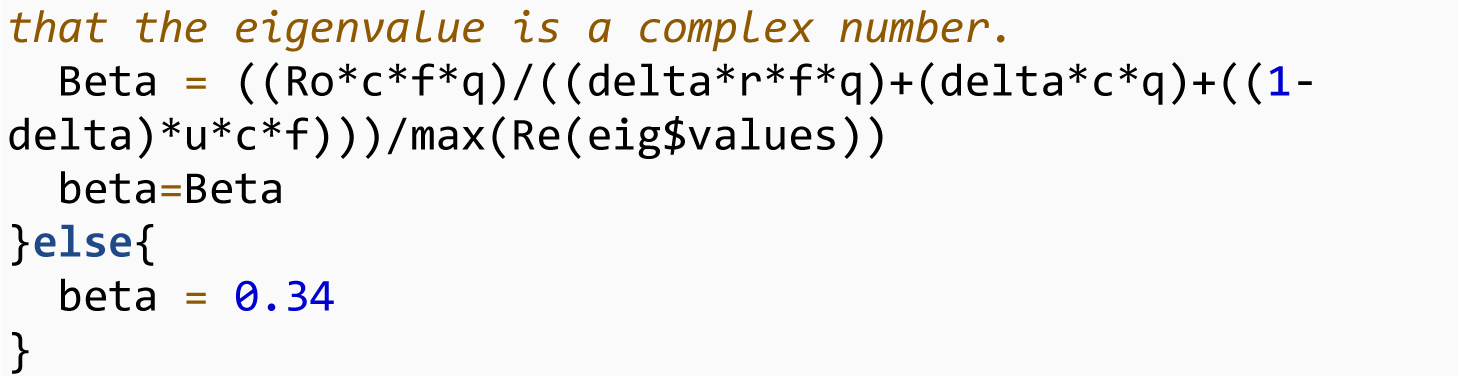

We defined the initial state conditions.

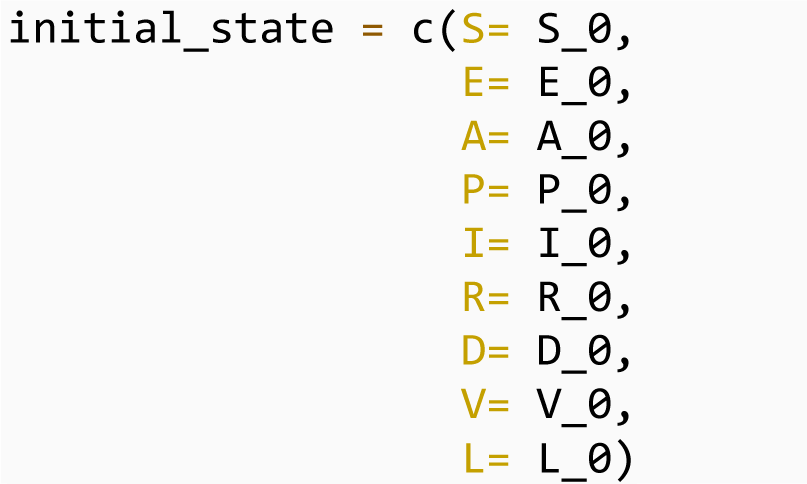

We created a vector for the parameters for initializing our model.

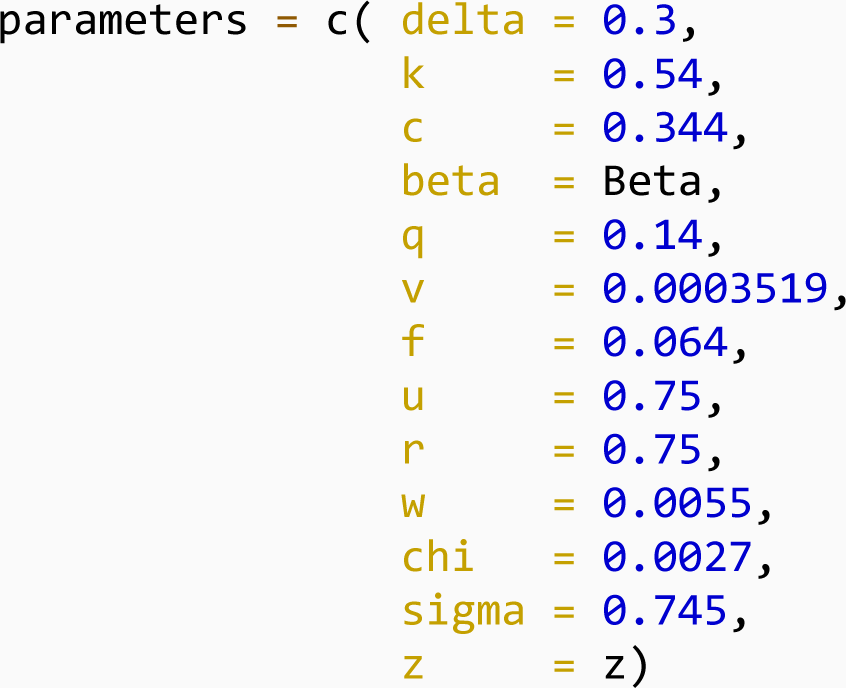

The model was simulated for 500 days to allow enough time for the second wave to emerge.

`time = seq (0, 500, 1)`

We solved the differential equations using the lsoda() function in the deSolve package:

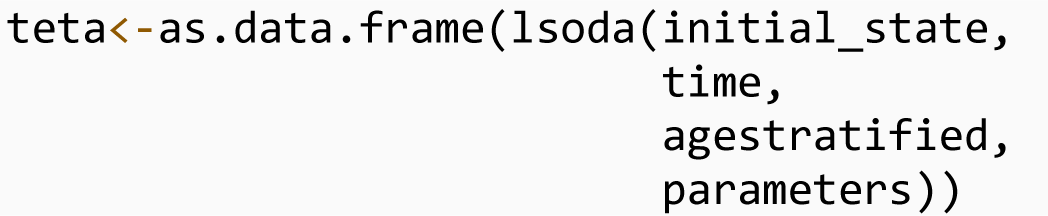

We converted the data into a long format using the melt() function in the reshape2 package:

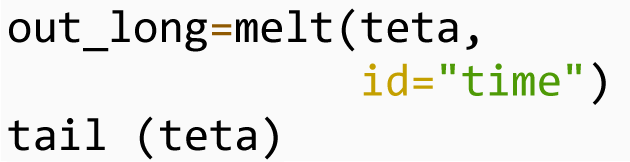

### Scenario analysis of the impact of vaccine optimization strategies

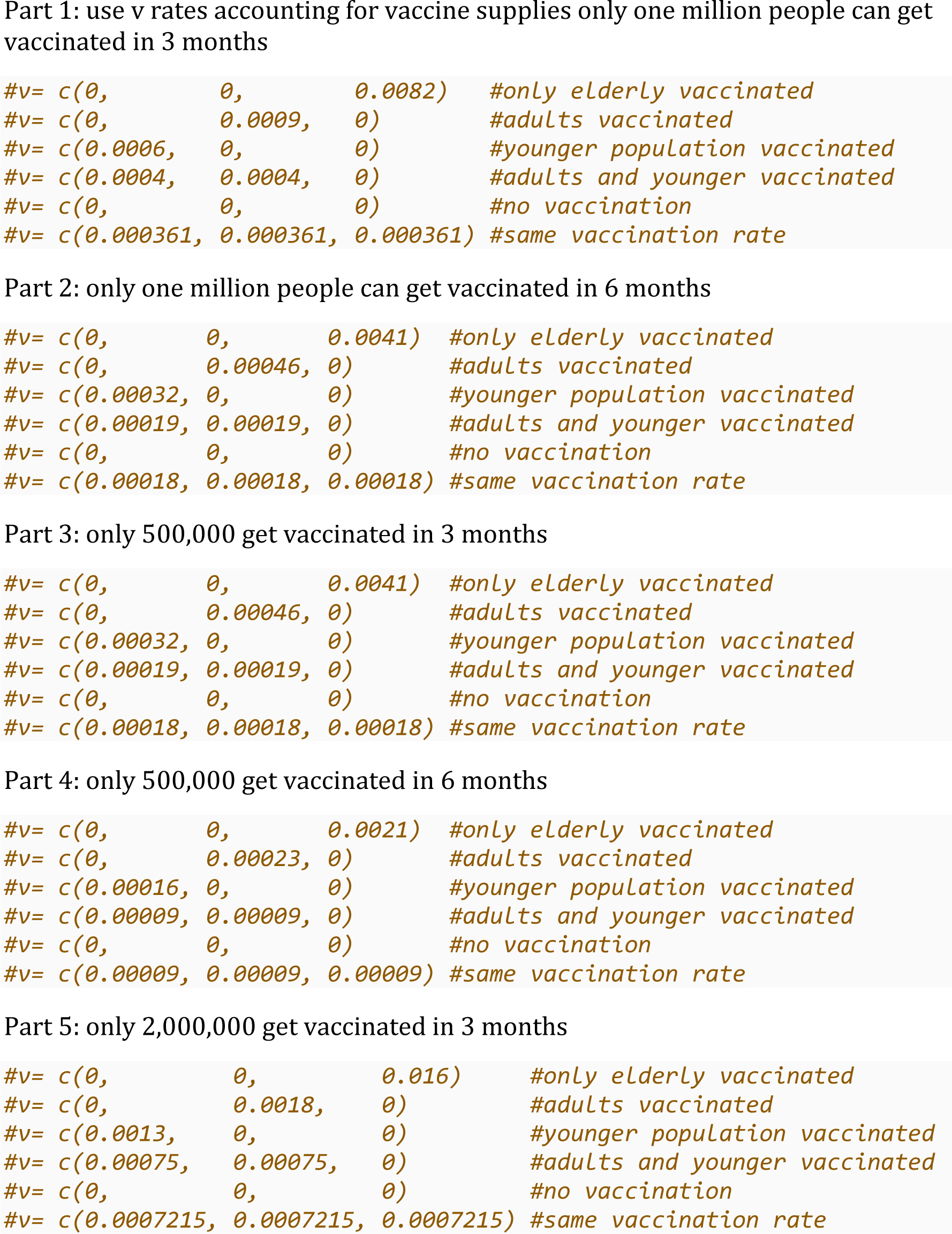
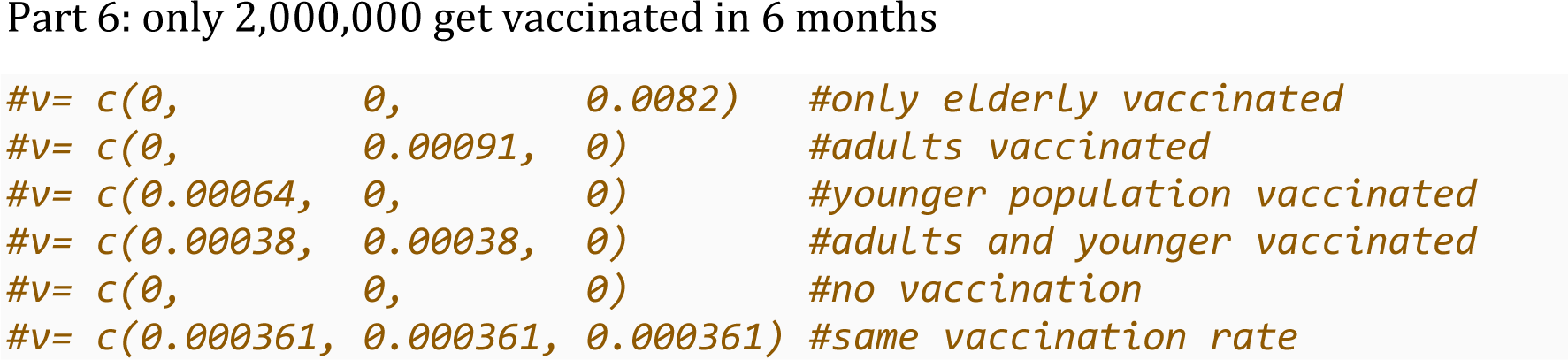

We defined parameter values for the simulation.

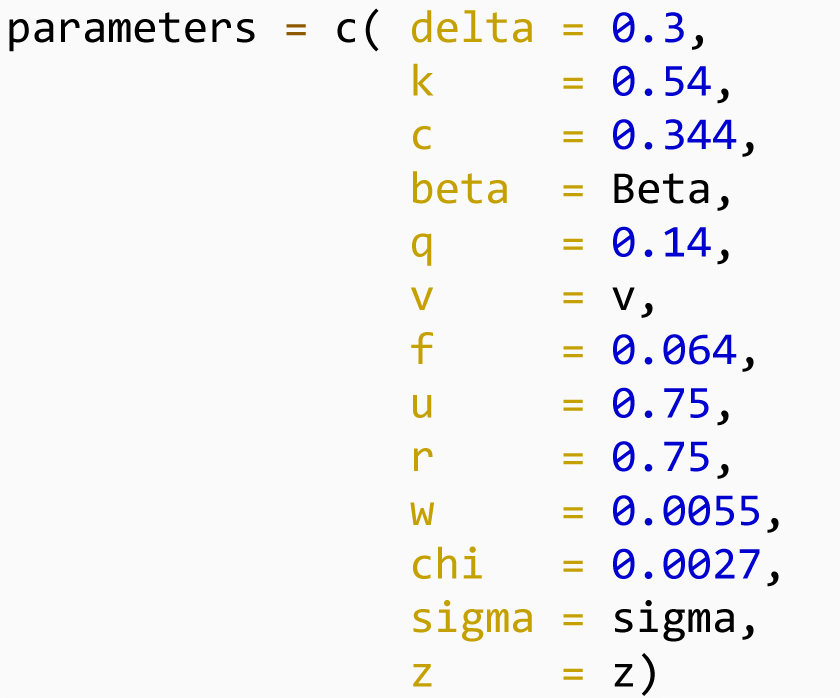

We solved the differential equations as:

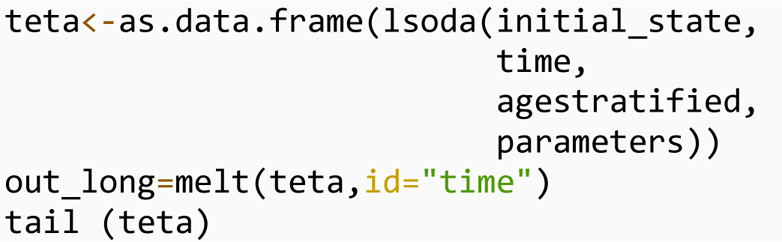

### Calculating the percentage of population with each outcome under the various vaccination scenarios

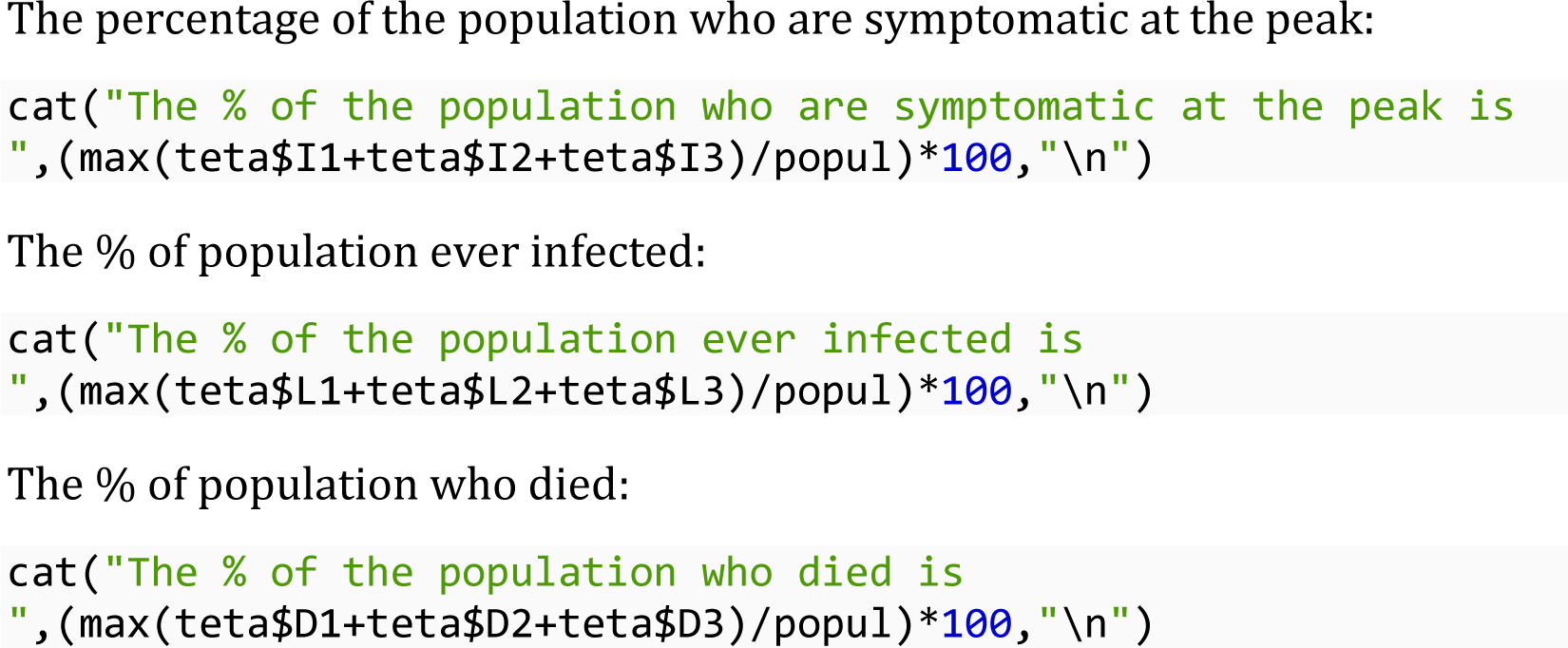

## Notes

### Funding Statement

This study did not receive any external funding.

## References

1. Cascella M, Rajnik M, Cuomo A, Dulebohn SC, Di Napoli R. Features, evaluation and treatment coronavirus (COVID-19). Statpearls [internet]: StatPearls Publishing; 2020.

2. Akande OW, Akande TM. COVID-19 pandemic: A global health burden. Niger Postgrad Med J. 2020 Jul-Sep;27(3):147-55.

3. Ghana Health Service. COVID-19 Ghana’s Outbreak Response Management Updates. 2020 [cited 2022 April 15, 2022]; Available from: https://ghanahealthservice.org/covid19/archive.php

4. Citi Newsroom. Omicron now most dominant COVID-19 variant in Ghana – WACCBIP. 2022.

5. Ofori SK, Schwind JS, Sullivan KL, Cowling BJ, Chowell G, Fung IC-H. Transmission dynamics of COVID-19 in Ghana and the impact of public health interventions. medRxiv : the preprint server for health sciences. 2021.

6. Quakyi NK, Agyemang Asante NA, Nartey YA, Bediako Y, Sam-Agudu NA. Ghana’s COVID-19 response: the Black Star can do even better. BMJ global health. 2021;6(3):e005569.

7. Alhassan RK, Aberese-Ako M, Doegah PT, Immurana M, Dalaba MA, Manyeh AK, et al. COVID-19 vaccine hesitancy among the adult population in Ghana: evidence from a pre- vaccination rollout survey. Tropical Medicine and Health. 2021 2021/12/16;49(1):96.

8. WHO Africa. More COVID-19 vaccines arrive in Ghana. 2021.

9. Ghana Health Service. COVID-19 Ghana’s Outbreak Response Management Updates. 2020 [cited 2022 February 21, 2022]; Available from: https://ghanahealthservice.org/covid19/archive.php

10. Botwe BO, Antwi WK, Adusei JA, Mayeden RN, Akudjedu TN, Sule SD. COVID-19 vaccine hesitancy concerns: Findings from a Ghana clinical radiography workforce survey. Radiography (Lond). 2021 Oct 8.

11. Quashie PK, Mutungi JK, Dzabeng F, Oduro-Mensah D, Opurum PC, Tapela K, et al. Trends of SARS-CoV-2 antibody prevalence in selected regions across Ghana. medRxiv. 2021.

12. Alagoz O, Sethi AK, Patterson BW, Churpek M, Alhanaee G, Scaria E, et al. The impact of vaccination to control COVID-19 burden in the United States: A simulation modeling approach. PloS one. 2021;16(7):e0254456.

13. McBryde ES, Meehan MT, Adegboye OA, Adekunle AI, Caldwell JM, Pak A, et al. Role of modelling in COVID-19 policy development. Paediatric respiratory reviews. 2020;35:57–60.

14. Kohli M, Maschio M, Becker D, Weinstein MC. The potential public health and economic value of a hypothetical COVID-19 vaccine in the United States: Use of cost-effectiveness modeling to inform vaccination prioritization. Vaccine. 2021;39(7):1157–64.

15. Moghadas SM, Vilches TN, Zhang K, Wells CR, Shoukat A, Singer BH, et al. The impact of vaccination on coronavirus disease 2019 (COVID-19) outbreaks in the United States. Clinical Infectious Diseases. 2021;73(12):2257–64.

16. Matrajt L, Eaton J, Leung T, Brown ER. Vaccine optimization for COVID-19: Who to vaccinate first? Science Advances. 2021;7(6):eabf1374.

17. Mumtaz GR, El-Jardali F, Jabbour M, Harb A, Abu-Raddad LJ, Makhoul M. Modeling the Impact of COVID-19 Vaccination in Lebanon: A Call to Speed-Up Vaccine Roll Out. Vaccines. 2021;9(7):697.

18. Bubar KM, Reinholt K, Kissler SM, Lipsitch M, Cobey S, Grad YH, et al. Model-informed COVID-19 vaccine prioritization strategies by age and serostatus. Science of the total environment. 2021;3711(6532):916-21.

19. University of British Columbia Department of Zoology. COVID-19 Models Lecture. 2020 [cited 2020 11/15/2020]; Available from: https://www.zoology.ubc.ca/~bio301/Bio301/Lectures/Lecture1/COVID_Models.pdf

20. Savvides C, Siegel R. Asymptomatic and presymptomatic transmission of SARS-CoV-2: A systematic review. medRxiv. 2020 Jun 17:2020.06.11.20129072.

21. Li Y, Shi J, Xia J, Duan J, Chen L, Yu X, et al. Asymptomatic and Symptomatic Patients With Non-severe Coronavirus Disease (COVID-19) Have Similar Clinical Features and Virological Courses: A Retrospective Single Center Study. Frontiers in microbiology. 2020;11:1570-.

22. Zou L, Ruan F, Huang M, Liang L, Huang H, Hong Z, et al. SARS-CoV-2 Viral Load in Upper Respiratory Specimens of Infected Patients. N Engl J Med. 2020 Mar 19;382(12):1177–9.

23. Zhao H, Lu X, Deng Y, Tang Y, Lu J. COVID-19: asymptomatic carrier transmission is an underestimated problem. Epidemiology and infection. 2020;148:e116-e.

24. Armachie J, Adom-Konadu A, Asiamah M, Dwomoh D. Impact of Partial lockdown on Time-Dependent Effective Reproduction Number of COVID-19 infection in Ghana: Application of change point analysis. Preprints. 2021.

25. Pearson CA, Silal SP, Li MW, Dushoff J, Bolker BM, Abbott S, et al. Bounding the levels of transmissibility & immune evasion of the Omicron variant in South Africa. Epidemics. 2021.

26. Bernal JL, Andrews N, Gower C, Gallagher E, Simmons R, Thelwall S, et al. Effectiveness of Covid-19 vaccines against the B. 1.617. 2 (Delta) variant. New England Journal of Medicine. 2021.

27. le Polain de Waroux O, Cohuet S, Ndazima D, Kucharski AJ, Juan-Giner A, Flasche S, et al. Characteristics of human encounters and social mixing patterns relevant to infectious diseases spread by close contact: a survey in Southwest Uganda. BMC Infectious Diseases. 2018 2018/04/11;18(1):172.

28. Kiti MC, Kinyanjui TM, Koech DC, Munywoki PK, Medley GF, Nokes DJ. Quantifying age-related rates of social contact using diaries in a rural coastal population of Kenya. PloS one. 2014;9(8):e104786.

29. Trentini F, Guzzetta G, Galli M, Zardini A, Manenti F, Putoto G, et al. Modeling the interplay between demography, social contact patterns, and SARS-CoV-2 transmission in the South West Shewa Zone of Oromia Region, Ethiopia. BMC medicine. 2021;19(1):1–13.

30. Central Intelligence Agency. The world factbook. 2020 [cited Jan 15, 2022]; Available from: https://www.cia.gov/the-world-factbook/

31. Anaadem P. Ghana: Population Hits 30.8 Million - Greater Accra Most Populous Region-- GSS. Ghana Today. 2021 September 22, 2021.

32. Soetaert KE, Petzoldt T, Setzer RW. Solving differential equations in R: package deSolve. Journal of statistical software. 2010;33.

33. Foy BH, Wahl B, Mehta K, Shet A, Menon GI, Britto C. Comparing COVID-19 vaccine allocation strategies in India: A mathematical modelling study. International journal of infectious diseases : IJID : official publication of the International Society for Infectious Diseases. 2021;103:431–8.

34. Ko Y, Lee J, Kim Y, Kwon D, Jung EJIJoER, Health P. COVID-19 vaccine priority strategy using a heterogenous transmission model based on maximum likelihood estimation in the Republic of Korea. 2021;18(12):6469.

35. Diarra M, Kebir A, Talla C, Barry A, Faye J, Louati D, et al. Non-pharmaceutical interventions and COVID-19 vaccination strategies in Senegal: a modelling study. BMJ global health. 2022;7(2):e007236.

36. Buckner JH, Chowell G, Springborn MR. Dynamic prioritization of COVID-19 vaccines when social distancing is limited for essential workers. Proceedings of the National Academy of Sciences. 2021;118(16).

37. Chapman LAC, Shukla P, Rodríguez-Barraquer I, Shete PB, León TM, Bibbins-Domingo K, et al. Risk factor targeting for vaccine prioritization during the COVID-19 pandemic. Scientific Reports. 2022 2022/02/23;12(1):3055.

38. Saadi N, Chi YL, Ghosh S, Eggo RM, McCarthy CV, Quaife M, et al. Models of COVID- 19 vaccine prioritisation: a systematic literature search and narrative review. BMC Med. 2021 Dec 1;19(1):318.

39. Acheampong T, Akorsikumah EA, Osae-Kwapong J, Khalid M, Appiah A, Amuasi JH. Examining Vaccine Hesitancy in Sub-Saharan Africa: A Survey of the Knowledge and Attitudes among Adults to Receive COVID-19 Vaccines in Ghana. Vaccines. 2021;9(8):814.

40. Nikolovski J, Koldijk M, Weverling GJ, Spertus J, Turakhia M, Saxon L, et al. Factors indicating intention to vaccinate with a COVID-19 vaccine among older U.S. adults. PloS one. 2021;16(5):e0251963-e.

41. Dembek ZF, Schwartz-Watjen KT, Swiatecka AL, Broadway KM, Hadeed SJ, Mothershead JL, et al. Coronavirus Disease 2019 on the Heels of Ebola Virus Disease in West Africa. Pathogens (Basel, Switzerland). 2021;10(10):1266.

42. Andrews N, Stowe J, Kirsebom F, Toffa S, Rickeard T, Gallagher E, et al. Covid-19 Vaccine Effectiveness against the Omicron (B.1.1.529) Variant. N Engl J Med. 2022 Mar 2.

43. Abbasi Z, Zamani I, Mehra AHA, Shafieirad M, Ibeas A. Optimal Control Design of Impulsive SQEIAR Epidemic Models with Application to COVID-19. Chaos, solitons, and fractals. 2020 2020-Oct;139:110054-.

44. Liu Z, Magal P, Seydi O, Webb G. A COVID-19 epidemic model with latency period. Infectious Disease Modelling. 2020;5:323–37.

45. Cai Q, Huang D, Ou P, Yu H, Zhu Z, Xia Z, et al. COVID-19 in a designated infectious diseases hospital outside Hubei Province, China. J Allergy. 2020.

46. Xing Y, Ni W, Wu Q, Li W, Li G, Tong J, et al. Prolonged presence of SARS-CoV-2 in feces of pediatric patients during the convalescent phase. medRxiv : the preprint server for health sciences. 2020.

47. Ma S, Zhang J, Zeng M, Yun Q, Guo W, Zheng Y, et al. Epidemiological parameters of coronavirus disease 2019: a pooled analysis of publicly reported individual data of 1155 cases from seven countries. MedRxiv : the preprint server for health sciences. 2020.

48. Byrne AW, McEvoy D, Collins AB, Hunt K, Casey M, Barber A, et al. Inferred duration of infectious period of SARS-CoV-2: rapid scoping review and analysis of available evidence for asymptomatic and symptomatic COVID-19 cases. BMJ open. 2020;10(8):e039856.

49. Tindale L, Coombe M, Stockdale JE, Garlock E, Lau WYV, Saraswat M, et al. Transmission interval estimates suggest pre-symptomatic spread of COVID-19. MedRxiv : the preprint server for health sciences. 2020.

50. Chen M, Li M, Hao Y, Liu Z, Hu L, Wang L. The introduction of population migration to SEIAR for COVID-19 epidemic modeling with an efficient intervention strategy. Information Fusion. 2020;64:252–8.

51. Buitrago-Garcia D, Egli-Gany D, Counotte MJ, Hossmann S, Imeri H, Ipekci AM, et al. Occurrence and transmission potential of asymptomatic and presymptomatic SARS-CoV-2 infections: A living systematic review and meta-analysis. PLoS medicine. 2020;17(9):e1003346-e.

52. Knoll MD, Wonodi C. Oxford-AstraZeneca COVID-19 vaccine efficacy. Lancet (London, England). 2021;397(10269):72–4.

53. Centers for Disease Control and Prevention. Pandemic Planning Scenarios. 2020 [cited 2020 November 12, 2020]; Available from: https://www.cdc.gov/coronavirus/2019-ncov/hcp/planning-scenarios.html

54. Iacobucci G. Covid-19: Protection from two doses of vaccine wanes within six months, data suggest. British Medical Journal Publishing Group; 2021.

55. Good MF, Hawkes MT. The Interaction of Natural and Vaccine-Induced Immunity with Social Distancing Predicts the Evolution of the COVID-19 Pandemic. Mbio. 2020;11(5).

56. Lawal Y. Africa’s low COVID-19 mortality rate: A paradox? International journal of infectious diseases : IJID : official publication of the International Society for Infectious Diseases. 2021;102:118–22.

## References

1. Liu R, Leung RK-k, Chen T, Zhang X, Chen F, Chen S, et al. The Effectiveness of Age- Specific Isolation Policies on Epidemics of Influenza A (H1N1) in a Large City in Central South China. PLoS One. 2015;10(7):e0132588.

2. Zhao Z-Y, Zhu Y-Z, Xu J-W, Hu S-X, Hu Q-Q, Lei Z, et al. A five-compartment model of age-specific transmissibility of SARS-CoV-2. Infectious Diseases of Poverty. 2020 2020/08/26;9(1):117.

3. Abbasi Z, Zamani I, Mehra AHA, Shafieirad M, Ibeas A. Optimal Control Design of Impulsive SQEIAR Epidemic Models with Application to COVID-19. Chaos, Solitons & Fractals. 2020 2020/10/01/;139:110054.

4. Liu Z, Magal P, Seydi O, Webb G. A COVID-19 epidemic model with latency period. Infectious Disease Modelling. 2020;5:323–37.

5. Chen M, Li M, Hao Y, Liu Z, Hu L, Wang L. The introduction of population migration to SEIAR for COVID-19 epidemic modeling with an efficient intervention strategy. Information Fusion. 2020;64:252–8.

6. Buitrago-Garcia D, Egli-Gany D, Counotte MJ, Hossmann S, Imeri H, Ipekci AM, et al. Occurrence and transmission potential of asymptomatic and presymptomatic SARS-CoV-2 infections: A living systematic review and meta-analysis. PLoS Med. 2020;17(9):e1003346.

7. Tindale L, Coombe M, Stockdale JE, Garlock E, Lau WYV, Saraswat M, et al. Transmission interval estimates suggest pre-symptomatic spread of COVID-19. medRxiv. 2020.

8. Cai Q, Huang D, Ou P, Yu H, Zhu Z, Xia Z, et al. COVID-19 in a designated infectious diseases hospital outside Hubei Province, China. Allergy. 2020;75(7):1742–52.

9. Byrne AW, McEvoy D, Collins AB, Hunt K, Casey M, Barber A, et al. Inferred duration of infectious period of SARS-CoV-2: rapid scoping review and analysis of available evidence for asymptomatic and symptomatic COVID-19 cases. BMJ open. 2020;10(8):e039856.

10. Thiruvengadam G, Ramanujam R, Marappa L. Modeling the recovery time of patients with coronavirus disease 2019 using an accelerated failure time model. Journal of International Medical Research. 2021;49(8):03000605211040263.

11. Xing Y-H, Ni W, Wu Q, Li W-J, Li G-J, Wang W-D, et al. Prolonged viral shedding in feces of pediatric patients with coronavirus disease 2019. Journal of microbiology, immunology infection. 2020;53(3):473–80.

12. University of British Columbia Department of Zoology. COVID-19 Models Lecture. 2020 [cited 2020 11/15/2020]; Available from: https://www.zoology.ubc.ca/~bio301/Bio301/Lectures/Lecture1/COVID_Models.pdf

13. Armachie J, Adom-Konadu A, Asiamah M, Dwomoh D. Impact of Partial lockdown on Time-Dependent Effective Reproduction Number of COVID-19 infection in Ghana: Application of change point analysis. Preprints. 2021.

14. Centers for Disease Control and Prevention. Pandemic Planning Scenarios. 2020 [cited 2020 November 12, 2020]; Available from: https://www.cdc.gov/coronavirus/2019-ncov/hcp/planning-scenarios.html

15. Knoll MD, Wonodi C. Oxford-AstraZeneca COVID-19 vaccine efficacy. Lancet. 2021;397(10269):72–4.

16. Good MF, Hawkes MT. The Interaction of Natural and Vaccine-Induced Immunity with Social Distancing Predicts the Evolution of the COVID-19 Pandemic. Mbio. 2020;11(5).

17. Keeling MJ, White PJ. Targeting vaccination against novel infections: risk, age and spatial structure for pandemic influenza in Great Britain. Journal of the Royal Society Interface. 2011;8(58):661–70.

18. Mary Eyram Ashinyo VD, Stephen Dajaan Dubik, Kingsley Ebenezer Amegah, Selorm Kutsoati, Ebenezer Oduro-Mensah, Peter Puplampu, Martha Gyansa-Lutterodt, Delese Mimi Darko, Kwame Ohene Buabeng, Anthony Ashinyo, Anthony Adofo Ofosu, Nyonuku Akosua Baddoo, Samuel Kaba Akoriyea, Francis Ofei, Patrick Kuma-Aboagye. Clinical characteristics, treatment regimen and duration of hospitalization among COVID-19 patients in Ghana: a retrospective cohort study. Pan African Medical Journal. 2020;2(PAMJ Special issue on COVID 19 in Africa).

19. Factbook CIA. The world factbook. See also: https://www.ciagov/library/publications/the-world-factbook. 2020.

20. Anderson RM, May RM. Infectious diseases of humans: dynamics and control: Oxford university press; 1992.

21. Towers S, Feng Z. Social contact patterns and control strategies for influenza in the elderly. Mathematical biosciences. 2012;240(2):241–9.

22. Towers S. SIR infectious disease model with age classes. 2012 [cited 2021 09/15/2021]; Available from: http://sherrytowers.com/2012/12/11/sir-model-with-age-classes/

23. le Polain de Waroux O, Cohuet S, Ndazima D, Kucharski AJ, Juan-Giner A, Flasche S, et al. Characteristics of human encounters and social mixing patterns relevant to infectious diseases spread by close contact: a survey in Southwest Uganda. BMC Infect Dis. 2018 2018/04/11;18(1):172.

24. Kiti MC, Kinyanjui TM, Koech DC, Munywoki PK, Medley GF, Nokes DJ. Quantifying age-related rates of social contact using diaries in a rural coastal population of Kenya. PLoS One. 2014;9(8):e104786.

25. Melegaro A, Jit M, Gay N, Zagheni E, Edmunds WJ. What types of contacts are important for the spread of infections? Using contact survey data to explore European mixing patterns. Epidemics. 2011;3(3-4):143–51.

26. Trentini F, Guzzetta G, Galli M, Zardini A, Manenti F, Putoto G, et al. Modeling the interplay between demography, social contact patterns, and SARS-CoV-2 transmission in the South West Shewa Zone of Oromia Region, Ethiopia. BMC medicine. 2021;19(1):1–13.

27. Melegaro A, Del Fava E, Poletti P, Merler S, Nyamukapa C, Williams J, et al. Social contact structures and time use patterns in the Manicaland Province of Zimbabwe. PLoS One. 2017;12(1):e0170459.

28. Odikro MA, Kenu E, Malm KL, Asiedu-Bekoe F, Noora CL, Frimpong J, et al. Epidemiology of COVID-19 outbreak in Ghana, 2020. Ghana Med J. 2020;54(4s):5-15.

29. Lawal Y. Africa’s low COVID-19 mortality rate: A paradox? Int J Infect Dis. 2021;102:118–22.

30. Soetaert KE, Petzoldt T, Setzer RW. Solving differential equations in R: package deSolve. Journal of statistical software. 2010;33.

31. Quashie PK, Mutungi JK, Dzabeng F, Oduro-Mensah D, Opurum PC, Tapela K, et al. Trends of SARS-CoV-2 antibody prevalence in selected regions across Ghana. medRxiv. 2021.

